# SC-MAPS: Image-based deep learning for bacterial identification and antibiotic-susceptibility testing

**DOI:** 10.64898/2026.10.23.26366325

**Authors:** Anuj Tiwari, Piers Turner, Conor Feehily, Stelios Chatzimichail, Nicole Stoesser, Christoffer Nellåker, Achillefs N. Kapanidis

**Author notes:** These authors contributed equally (Anuj Tiwari, Piers Turner).

## Abstract

Rapid bacterial identification could help guide antimicrobial susceptibility testing (AST), but image-based classification must generalise across bacterial strains. We developed SC-MAPS, a multichannel microscopy dataset comprising 916,435 single cells from 13 bacterial species, with independent strains reserved for evaluation. Combining phase-contrast and fluorescence information using deep learning achieved 65.4% single-cell species-classification accuracy on held-out strains. To assess whether imperfect species predictions retained useful information for downstream testing, we defined eight antimicrobial-resistance-informed AST-routing groups. Pooling species probabilities increased routing accuracy to 77.1%, rising to 91.1% when predictions from five cells were combined and 96.2% with 20 cells. Synthetic mixtures revealed limitations in minority-species recovery, while exploratory analysis of unseen organisms demonstrated the potential for out-of-distribution detection. SC-MAPS provides a framework for investigating transferable bacterial phenotypes and developing image-based identification and AST-routing strategies.

## Introduction

Bacterial infections remain a major cause of morbidity and mortality worldwide, with their clinical burden increasingly compounded by antimicrobial resistance (AMR)^1,2^. Effective treatment depends not only on identifying the causative organism, but on doing so rapidly enough to guide appropriate antimicrobial susceptibility testing (AST) and reduce the period during which treatment must be selected empirically. Routine clinical microbiology still commonly relies on culture or pre-culture to generate sufficient bacterial biomass before identification using biochemical, molecular or proteomic approaches, including PCR and matrix-assisted laser desorption/ionisation time-of-flight mass spectrometry (MALDI-TOF MS)^3–6^. Although these methods are highly effective, conventional identification and susceptibility workflows can require many hours to days before actionable information becomes available^7,8^. Therefore, there is considerable interest in approaches that can extract useful phenotypic information from much smaller numbers of cells over shorter timeframes.

Importantly, the clinically relevant question is not necessarily limited to whether every bacterium can immediately be assigned to its exact species. Early information that distinguishes organisms associated with different downstream antimicrobial-testing strategies could potentially help guide the selection of an initial AST pathway.

Microscopy offers a potential route towards such early phenotypic information because bacterial morphology and cellular organisation can be examined directly at single-cell resolution. Conventional morphology alone has limited discriminatory power, as phylogenetically distinct bacteria can share similar dimensions and shapes, while cells of the same species can display substantial phenotypic variation^9^. Deep learning provides a means of extracting more complex image features that are difficult to capture using predefined morphological measurements^10,11^. Previous work has demonstrated the potential of this approach across increasingly sophisticated imaging tasks. Deep-learning analysis has been applied to bacterial colonies and gram-stained blood-culture images^12,13^, while single-cell approaches have shown that bacterial morphology can contain information relevant to species identity, physiological state and antimicrobial response^14–17^. Recent work further indicates that morphological differences can be associated with antimicrobial-resistance phenotypes even in the absence of antibiotic exposure^18^.

More information-rich techniques, including hyperspectral microscopy^19–22^, coherent imaging^23^, quantitative-phase microscopy^24^ and Raman spectroscopy^25–27^, have further demonstrated that individual bacterial cells contain phenotypic signatures capable of supporting classification. For example, quantitative-phase imaging achieved 82.5% species-identification accuracy from a single three-dimensional refractive-index tomogram of an individual bacterial cell or small cluster across 19 bloodstream-infection-associated species, increasing to 99.9% when predictions from seven independently measured cells or clusters were combined ^24^. This distinction is important for diagnostic applications: individual bacterial cells are heterogeneous and may be difficult to classify reliably in isolation, whereas evidence accumulated across a small population can provide substantially more robust sample-level information^28,29^.

The shift towards single-cell bacterial phenotyping has also been enabled by advances in image segmentation^30^. Deep-learning-based tools such as DeLTA^31^, MiSiC^32^, Omnipose^33^ and Cellpose^34^ can identify individual bacteria across crowded fields and variable cell morphologies, allowing image information to be analysed at the level of discrete cells rather than whole fields of view. This is particularly important for bacteria, where small cell size, close spatial packing and substantial morphological variation can complicate segmentation. Reliable single-cell extraction preserves phenotypic heterogeneity that may otherwise be obscured by population averaging and provides the basis for linking morphology, fluorescence sub-cellular distribution, and intracellular organisation to downstream classification.

However, high reported classification accuracy does not necessarily imply that a model will generalise to a previously unseen strain of the same bacterial species. This is highly relevant to single-cell datasets, where thousands of cells may originate from only a small number of strains, cultures or experimental preparations. Strain-to-strain variation introduces additional phenotypic heterogeneity within a species, meaning that models trained on a limited number of strains may learn strain-associated features that do not transfer to other members of the same species. Moreover, cells originating from the same culture or experimental unit share biological history and technical characteristics and therefore cannot be considered fully independent observations. If such cells are distributed between training and test sets, models may learn preparation- or experiment-associated features rather than phenotypes that generalise to independent material. Hallström et al. reported 99.55% mean accuracy in a controlled four-species phase-contrast time-lapse experiment ^35^, whereas extension to seven species and evaluation in a separate unseen mixed-species experiment yielded 93.5% precision and 94.7% recall ^36^. Similarly, Li et al. reported reduced single-cell Raman classification when biological replicates were withheld ^27^. Comparable distribution shifts have also been observed beyond microscopy: Wang et al. reported a decrease in median AUC for MALDI-TOF-based AMR prediction from 0.94 under internal evaluation to 0.55 on temporally external data ^37^. These studies illustrate that the number of cells analysed and the biological diversity represented by those cells are fundamentally different properties of a dataset. Large-scale single-cell resources therefore need to preserve both strain and experimental provenance and incorporate strain-level holdout evaluation to determine whether learned phenotypes transfer beyond the bacterial material used for model development.

A further opportunity lies in combining complementary phenotypic information from the same cell. Many high-performing bacterial classification approaches derive rich information from a single specialised measurement, including quantitative-phase, hyperspectral or Raman imaging. In contrast, conventional transmitted-light and fluorescence microscopy can provide several biologically distinct views of the same bacterium, including overall morphology, nucleoid organisation, membrane structure and cell-envelope-associated staining. Bacterial membrane and surface architecture are themselves heterogeneous phenotypes that vary between organisms and physiological states^38^. Recent multi-target fluorescence imaging of clinical bacterial isolates further supports the ability of complementary intracellular and cell-wall-associated signals to contribute to species discrimination^39^. However, it is unclear whether these complementary phenotypes can be integrated systematically at large scale while preserving single-cell correspondence, strain and experimental origin, and robust evaluation across independently held-out strains and experimental preparations. Addressing this gap requires datasets that are not only large in cell number, but also multimodal and structured around biologically meaningful experimental units.

We therefore developed SC-MAPS (Single-Cell Multi-channel Analysis for antibiotic-Panel and Species classification), a large-scale, spatially matched multichannel single-cell microscopy resource comprising 916,435 cells from 13 clinically relevant Gram-positive (n = 4) and Gram-negative (n = 9) bacterial species. SC-MAPS combines phase-contrast and bright-field imaging of cellular morphology with DAPI staining of nucleic acids and nucleoid organisation, WGA-AF647 labelling of cell-envelope-associated glycoconjugates, and FM1-43 staining of the cell membrane, allowing complementary cellular phenotypes to be examined individually and in combination for bacterial classification.

Model development and final testing were separated at the strain level: two to three strains per species were used for model development, while an additional strain was reserved exclusively for independent holdout evaluation. This allowed us to test whether image-derived phenotypes generalised to previously unseen strains of the same species. Multichannel feature fusion improved species identification, while broader AMR-informed AST-routing classification was more robust than exact species classification. Combining predictions across multiple cells further improved both species- and AST-routing performance.

## Results

### SC-MAPS captures large-scale, multimodal and biologically heterogeneous single-cell bacterial phenotypes

The final SC-MAPS dataset comprised 916,435 individually segmented bacterial cells from 13 species, with each cell represented by spatially matched phase-contrast, bright-field (BF), DAPI, WGA-AF647 and FM1-43 images. Representative unmasked single-cell crops illustrate the correspondence between cellular morphology and the three fluorescence signals within the same bacterium (**Figure 1A**). All 13 species contributed tens of thousands of cells, ranging from 26,029 cells for SEPI to 140,938 for SLUG (**Figure 1B**).

**Figure 1:**
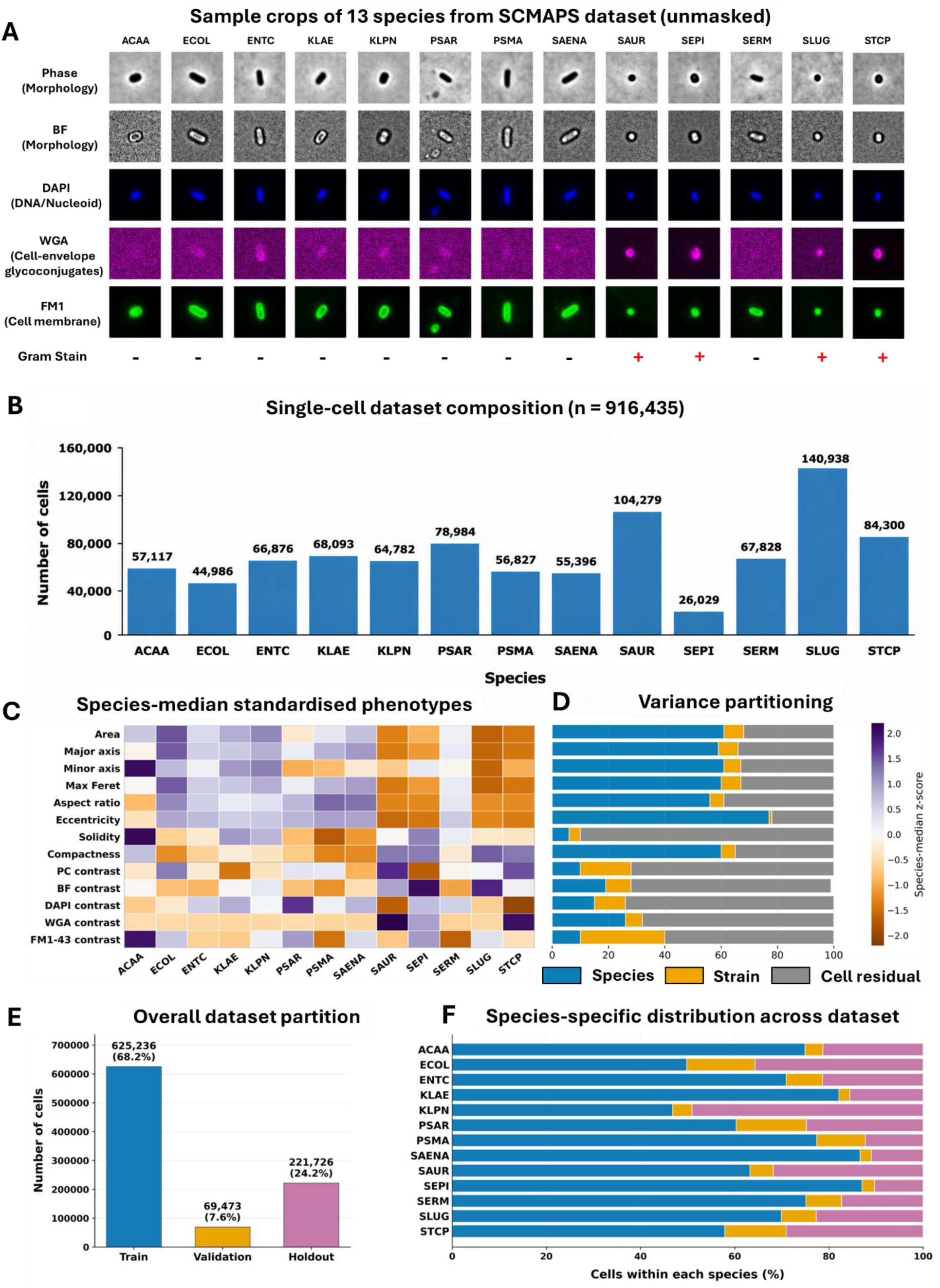
Composition and phenotypic structure of the SC-MAPS dataset. **(A)** Representative unmasked single-cell crops from 13 bacterial species imaged using phase contrast, bright field (BF), DAPI, WGA-AF647 and FM1-43. The imaging channels provide complementary information on cellular morphology, nucleoid organisation, cell-envelope-associated glycoconjugates and membrane structure. Scale bars, 2.5 µm **(B)** Distribution of the 916,435 segmented cells across the 13 species. **(C)** Standardised species-median phenotypes, comprising eight morphological measurements and five channel-contrast features. Values represent z-scores calculated across species for each phenotype. **(D)** Variance partitioning of the same phenotypes into components attributable to species identity, strain within species and residual cell-to-cell variation. **(E)** Overall numbers of cells assigned to the training, validation and independent holdout partitions. **(F)** Species-specific proportions of cells across the same dataset partitions. The holdout partition comprised an independently held-out strain for each species. Species abbreviations: **ACAA**, *Acinetobacter baumannii*; **ECOL**, *Escherichia coli*; **ENTC**, *Enterobacter cloacae*; **KLAE**, *Klebsiella aerogenes*; **KLPN**, *Klebsiella pneumoniae*; **PSAR**, *Pseudomonas aeruginosa*; **PSMA**, *Stenotrophomonas maltophilia*; **SAENA**, *Salmonella enterica*; **SAUR**, *Staphylococcus aureus*; **SEPI**, *Staphylococcus epidermidis*; **SERM**, *Serratia marcescens*; **SLU9**, *Staphylococcus lugdunensis*; and **STCP**, *Staphylococcus capitis*.

SC-MAPS also captured substantial phenotypic diversity within and between species. Quantitative single-cell measurements showed species-associated differences in cell area, major- and minor-axis lengths, maximum Feret diameter, aspect ratio, eccentricity, solidity and compactness (**Figure 1C**). Variance partitioning was used to estimate how much of the observed variation in each feature was associated with species identity, differences between strains within species, or residual variation among individual cells within each species–strain population. Species identity accounted for 60.9% of variation in cell area, 59.1% in major-axis length, 60.4% in maximum Feret diameter and 76.7% in eccentricity, while substantial residual variation remained among individual cells within species–strain populations (**Figure 1D**). Imaging-derived measurements showed greater strain/preparation- and cell-level variation. Species accounted for 9.3%, 19.4%, 14.9%, 25.5% and 10.5% of the variation in phase-contrast, BF, DAPI, WGA-AF647 and FM1-43 contrast, respectively. For these imaging-derived features, the strain/preparation component may also include acquisition-associated variation because fluorescence exposure times were not fixed across acquisitions. This component was particularly large for FM1-43, for which 29.6% of total contrast variance was attributable to strain/preparation differences within species (**Figure 1D**). Details of feature standardisation and the nested species–strain-within-species–cell variance decomposition are provided in Supplementary Methods Section 4.

The dataset was partitioned at the strain rather than individual-cell level (for exact partition sizes, see **Supplementary Table 3**; for the resulting species–strain cell distribution, see **Supplementary Figure S2**). Strains corresponding to Plates 2–4 were used for model development, contributing 625,236 training cells and 69,473 validation cells, whereas all 221,726 cells from the strains corresponding to Plate 1 were reserved for independent holdout evaluation (**Figure 1E**). This separation was retained across species (**Figure 1F**), ensuring that final performance was measured on cells from a strain that had not been encountered during model development.

### Species-level predictions preserve clinically relevant information for AST routing

We first established how effectively the multichannel phenotypes represented in SC-MAPS supported single-cell species classification. Because the holdout strain for each species was excluded entirely from model development, holdout performance measures generalisation to a previously unseen strain rather than classification of additional cells from strains represented during training. This strain-level separation therefore provides a more stringent assessment of within-species generalisation than random cell-level partitioning. Detailed benchmarking of individual imaging channels, probability-level ensembles and feature-fusion strategies is provided in **Supplementary Tables 4–6** and **Supplementary Methods Sections 5.3 – 5.5**.

### Species-level performance

Across DenseNet121, ResNet18 and ResNet34, classification performance depended more strongly on imaging modality than on CNN architecture. Among individual channels, phase contrast showed the strongest cross-strain generalisation to the independent holdout, reaching 54.8% accuracy with DenseNet121 (**Supplementary Table 4**). DAPI was the next strongest individual modality, whereas WGA-AF647 and FM1-43 showed more species-dependent performance and BF was consistently the weakest input. Combining information across channels improved performance: averaging class probabilities generated by independently trained DAPI, WGA-AF647, FM1-43 and phase-contrast models increased DenseNet121 holdout accuracy to 61.2% (**Supplementary Table 5**), while feature-level fusion further increased independent-strain holdout accuracy to 65.4% (**Supplementary Table 6**). Differences between CNN architectures and between concatenation and attention-based fusion were comparatively modest, and the DenseNet121 concatenation model was therefore used for subsequent analyses. A specialist classifier restricted to *Enterobacter cloacae* (ENTC), *Klebsiella aerogenes* (KLAE), *Klebsiella pneumoniae* (KLPN) and *Serratia marcescens* (SERM) did not improve generalisation to the independently held-out strains (51.5% accuracy; macro-F1 = 0.500; **Supplementary Methods Section 5.6** and **Supplementary Fig. S4**).

### AST-routing performance

Since exact species identification remained imperfect under independent strain-level evaluation, we next asked whether these predictions nevertheless retained information relevant to downstream antimicrobial susceptibility testing. The 13 species were grouped a priori into eight AMR-informed AST-routing groups representing broad differences in initial susceptibility-testing requirements: routine Enterobacterales, AmpC/caution Enterobacterales, Salmonella, Pseudomonas, Acinetobacter, Stenotrophomonas, an *S. aureus*-like group and coagulase-negative staphylococci (CoNS). The grouping strategy reflected species-specific differences in initial AST requirements and resistance-associated interpretive considerations. In particular, routine Enterobacterales were distinguished from organisms requiring additional AmpC-related interpretive caution, while Salmonella, Pseudomonas, Acinetobacter and Stenotrophomonas were retained as separate categories because of their distinct susceptibility-testing considerations. Among staphylococci, S. lugdunensis was grouped with S. aureus to reflect its comparatively invasive clinical behaviour, distinguishing these organisms from other coagulase-negative staphylococci. Consequently, some species-level misclassifications could still preserve the appropriate initial AST-routing category. These groups were therefore used for routing rather than as predictions of antimicrobial susceptibility, MIC, resistance mechanism or treatment response. Their species composition and clinical rationale are provided in **Supplementary Table 2**.

Species predictions were first converted to routing predictions by mapping the highest-probability species to its corresponding AST-routing group. This increased holdout accuracy from 65.4% at exact species level to 76.9% at the AST-routing-group level (**Figure 2A, B**). Of the 76,818 cells misclassified at species level, 26,530 (34.5%) nevertheless remained within the correct AST-routing group. Errors were not distributed uniformly across groups, with prominent confusion among the Enterobacterales routing groups and between the *S. aureus*-like and CoNS groups, while Stenotrophomonas also showed lower separability (**Figure 2A**). Thus, broader routing performance partly reflected biologically structured species-level errors rather than a uniform improvement across all organisms.

**Figure 2:**
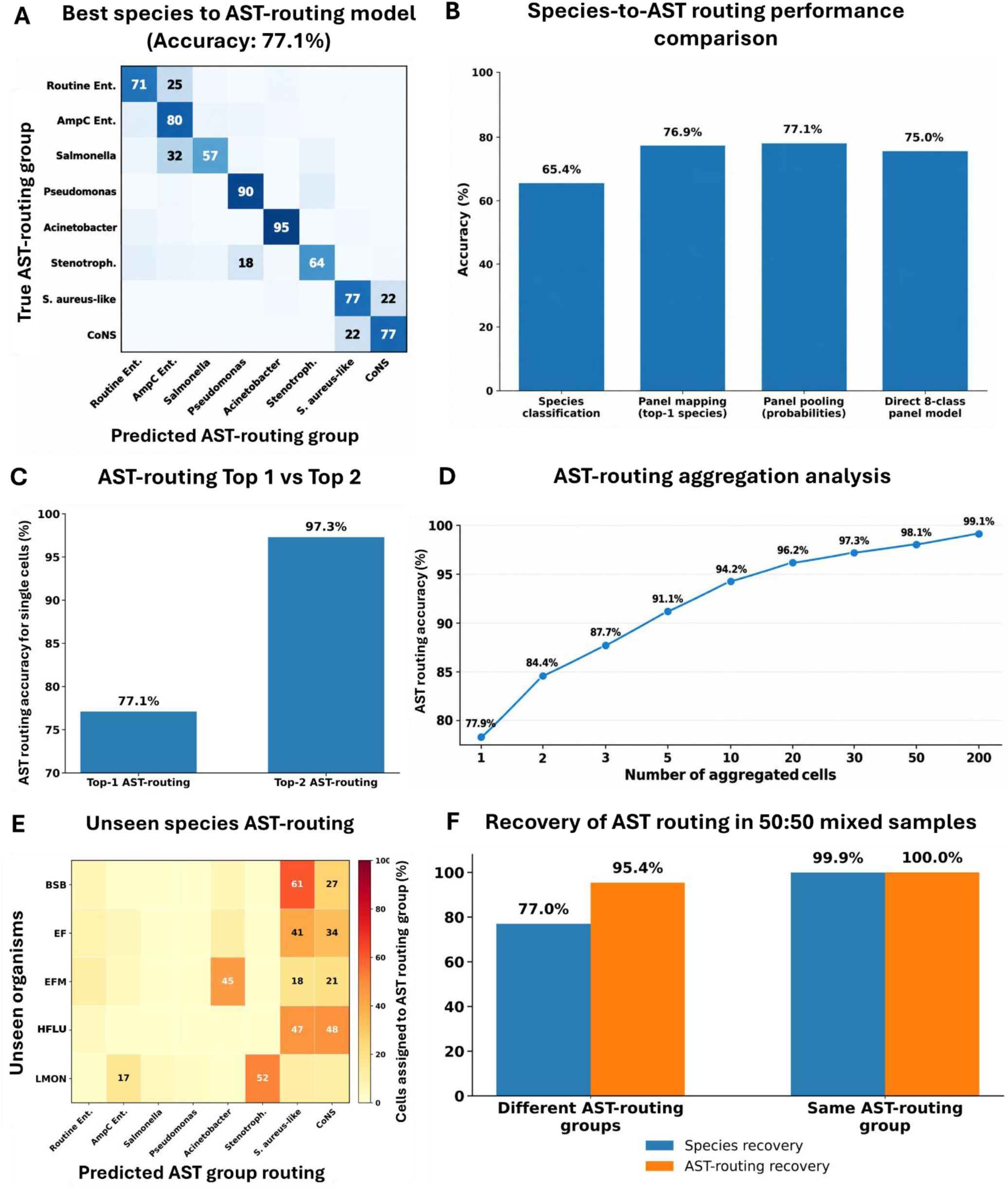
AST-routing from species-level predictions. **(A)** Row-normalised confusion matrix for eight AST-routing panels obtained by pooling species-level probabilities. **(B)** Comparison of species classification, top-1 species-to-panel mapping, probability-based pooling and direct eight-class AST grouping classification. **(C)** Top-1 and top-2 panel accuracy using pooled species probabilities. **(D)** Improvement in AST routing accuracy with increasing numbers of aggregated cells from the same sample. **(E)** AST routing of species excluded from model training, showing structured assignment of unseen organisms across the eight AST categories. **(F)** Recovery of constituent species and AST-routing groups in balanced 50:50 synthetic two-species populations. For 72 species pairs belonging to different AST-routing groups, recovery required both constituent species or both corresponding routing groups to appear among the two highest aggregate predictions. For six pairs sharing an AST-routing group, species recovery required identification of both constituent species, whereas AST-routing recovery required identification of their shared group.

To determine whether this improvement could arise simply from grouping 13 species into fewer classes, we compared the observed routing performance with a group-size-matched randomisation in which species were reassigned to eight groups while preserving the observed routing-group sizes. Top-1 species-to-routing mapping achieved 76.9% accuracy compared with a randomised mean of 68.0% (empirical *P* = 0.0087). We next retained the complete species-probability distribution and summed probabilities for species belonging to the same routing group. This probability-pooling approach produced the strongest single-cell routing performance, reaching 77.1% accuracy and a macro-F1 of 74.8% (**Figure 2A, B**), compared with a group-size-matched randomised mean accuracy of 68.1% (*P* = 0.0115).

Training DenseNet121 directly on the eight AST-routing groups did not improve classification accuracy, achieving approximately 75.0% compared with 77.1% following species-level training and probability pooling (**Figure 2B**). Sample-level bootstrap analysis yielded 95% confidence intervals of 75.5–78.8% for species-derived probability pooling and 73.0–76.9% for direct eight-class classification. These marginal intervals overlapped and did not independently establish a statistically significant difference between the two approaches. The correct AST-routing group was also contained within the two highest-probability pooled routing predictions for 97.3% of individual cells (**Figure 2C**), compared with a group-size-matched randomised mean of 86.8% (*P* = 0.0005). Together, these results indicate that species-level predictions retained routing information beyond that expected simply from collapsing species into larger groups. A complete comparison of species-derived and directly trained AST-routing strategies is provided in **Supplementary Table 7** and **Supplementary Methods Section 6.1-6.4.**

Classification difficulty varied across the 134-holdout sample/image-acquisition units. Median within-sample species accuracy was 64.7%, increasing to 78.0% at the AST-routing level. Sample-level performance was highly concordant between the concatenation and SE-attention fusion models at both species level (Spearman ρ = 0.964) and routing-group level (ρ = 0.945), indicating that samples that were difficult for one fusion strategy tended also to be difficult for the other. Of these 134 samples, 120 contained at least 200 segmented cells and were retained for multi-cell aggregation. The definition of a holdout sample and full within-sample analysis are provided in **Supplementary Methods Section 6.5**. We therefore tested whether combining predictions from multiple cells from the same sample could improve classification.

### Identification based on sets of single cells

Species-probability distributions from increasing numbers of randomly selected cells from the same sample were averaged to generate a single sample-level prediction. Species-level accuracy increased from 66.3% with one sample-balanced cell to 80.5% with five cells, 84.7% with ten, 86.5% with twenty and 89.0% with 200 cells. The one-cell value differs slightly from the overall 65.4% single-cell accuracy because each sample contributed equally to the aggregation analysis, rather than samples containing more segmented cells contributing proportionally more predictions. The effect was greater at the AST-routing level, where accuracy increased from 77.9% with one cell to 91.1% with five, 94.2% with ten and 96.2% with twenty cells, reaching 99.1% with 200 cells (**Figure 2D**). Thus, combining predictions across small numbers of cells substantially improved both species identification and AST-routing performance on strains absent from model development, with the largest gains occurring within the first 10–20 cells. Full aggregation procedures, including repeated subsampling and probability averaging, are provided in **Supplementary Methods Section 6.6**.

### Evaluating unseen species

Whereas the primary holdout tested generalisation to unseen strains of species represented during training, we next examined model behaviour for organisms belonging to species entirely absent from model development. Five unseen species were evaluated: *Streptococcus agalactiae* (BSB), *Enterococcus faecalis* (EF), *Enterococcus faecium* (EFM), *Listeria monocytogenes* (LMON) and *Haemophilus inffuenzae* (HFLU); corresponding strain identities are provided in **Supplementary Table 8**. When their forced species probabilities were pooled into the eight AST-routing groups, assignments were non-uniform (**Figure 2E**). BSB predictions were concentrated mainly within the *S. aureus*-like and CoNS groups, EFM preferentially mapped towards Acinetobacter, and LMON towards Stenotrophomonas. These structured assignments indicate that unseen organisms can share image-derived phenotypes with classes represented during training, but they do **not** demonstrate generalisation of AST routing to unseen species. Indeed, because the model is closed-set, every unseen cell is necessarily assigned to one of the learned classes, and some of these associations are not clinically meaningful. The results therefore illustrate both phenotypic relationships captured by the learned representation and the limitation of applying a closed-set classifier to organisms outside its training distribution. Unseen-organism cell numbers, forced-probability calculations and AST-routing analysis are described in **Supplementary Methods Section 7.1**.

### AST-routing recovery in synthetic mixtures

Balanced 50:50 synthetic mixtures were evaluated across all 78 unordered pairs of the 13 SC-MAPS species. Of these, 72 species pairs belonged to different AST-routing groups and six pairs belonged to the same routing group. For mixtures containing species from different routing groups, both constituent species were recovered among the two highest aggregate species predictions in 77.0% of mixtures, whereas both corresponding AST-routing groups were recovered in 95.4% (**Figure 2F**). To determine whether this increase could be explained simply by grouping species into broader categories, we repeated the analysis across 2,000 group-size-matched randomisations in which the two constituent species remained assigned to different groups. Mean recovery under this randomised null was 78.9%, significantly below the observed 95.4% (empirical *P* = 0.0015). Because the same- and different-routing-group analyses comprise different species-pair sets, the 99.9–100% recovery observed for same-routing-group pairs was treated descriptively and was not interpreted as evidence that shared routing-group membership itself improves recoverability. Synthetic-mixture generation, constituent-recovery definitions and group-size-matched randomisation procedures are described in **Supplementary Sections G.1–G.2**.

### Species-level analysis reveals structured phenotypic similarities and classification errors

We next examined how species were distinguished by the best-performing four-channel DenseNet121 concatenation model. The row-normalised confusion matrix showed substantial variation in recall across the 13 species (**Figure 3A**). ACAA, ECOL, PSAR and SLUG were among the best-resolved species, with recalls of approximately 82–95%, whereas recurrent errors occurred among several Enterobacterales and staphylococci. Species-level errors were therefore concentrated among particular classes rather than distributed uniformly across the dataset.

**Figure 3:**
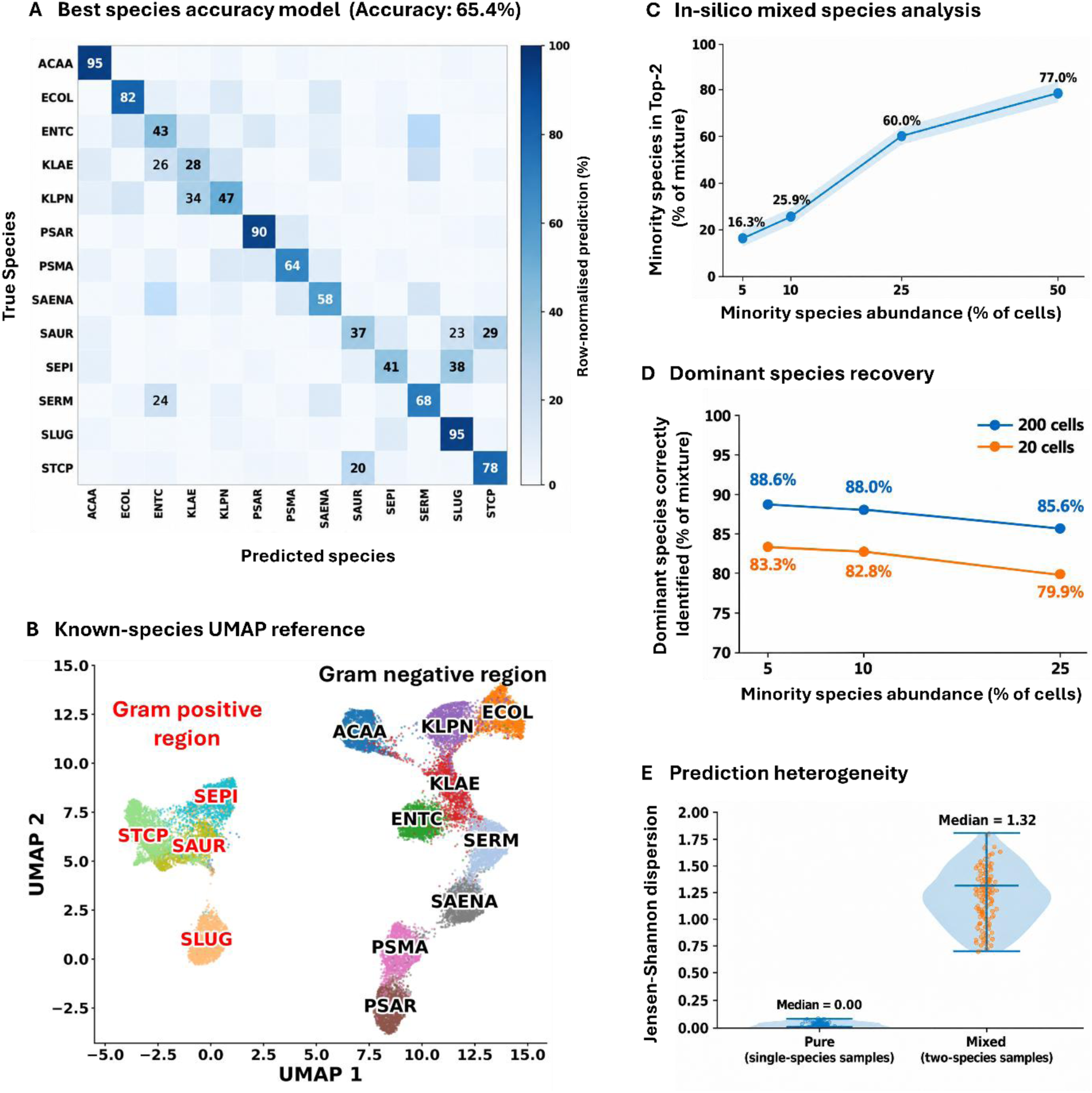
Species classification, learned feature organisation and behaviour in synthetic mixed populations. (A) Row-normalised confusion matrix for the four-channel DenseNet121 concatenation model evaluated on the independent-strain holdout. (B) UMAP of the learned feature representation for the 13 known SC-MAPS species. Species are shown in distinct colours, with Gram-positive species labels highlighted in red. (C) Minority-species recovery among the two highest-ranked aggregate species predictions in 200-cell synthetic two-species populations comprising species from different AST-routing groups (72 unordered species pairs). At 50:50 abundance, recovery required both constituent species to appear among the two highest aggregate predictions. (D) Dominant-species recovery in 20- and 200-cell synthetic mixtures across increasing minority-species abundance. (E) Generalized Jensen–Shannon dispersion of cell-level species-prediction distributions in pure and synthetic mixed 20-cell populations, showing substantially greater prediction heterogeneity in mixed populations.

To visualise the organisation of the learned representation, DenseNet121 feature vectors from the 13 known SC-MAPS species were projected using Uniform Manifold Approximation and Projection (UMAP; **Figure 3B**). Many species occupied distinct regions of the embedding, while overlap remained between several classes that were also difficult to distinguish in the confusion matrix. A broader separation was evident between the Gram-positive and Gram-negative species represented in the training set. UMAP was used here as a qualitative visualisation of learned feature organisation rather than as a quantitative measure of inter-species distance.

### Single-cell prediction heterogeneity reveals mixed populations and limits indiscriminate aggregation

Because combining predictions across cells substantially improved classification in homogeneous samples, we next examined how aggregation behaved when populations contained more than one species. Synthetic two-species populations were constructed from cells belonging to the independently held-out strains at defined relative abundances. In 200-cell populations, minority abundances of 5%, 10%, 25% and 50% corresponded to 10, 20, 50 and 100 cells from the less abundant constituent, respectively. Cell-level species probabilities were averaged across each population, and recovery was defined by whether the constituent species appeared among the two highest-probability aggregate species predictions.

Minority-species recovery increased from 16.3% of populations at 5% abundance to 25.9% at 10% and 60.0% at 25%; in balanced 50:50 populations, both constituent species were recovered among the two highest aggregate predictions in 77.0% of mixtures (**Figure 3C**). Thus, the 77.0% value represents successful recovery of both constituent species at the population level rather than correct classification of 77 of the 100 minority-species cells. In contrast, recovery of the dominant species remained comparatively stable across asymmetric mixtures (**Figure 3D**), indicating that population averaging preferentially preserves the dominant organism while potentially obscuring a less abundant constituent.

Mixed populations also showed greater disagreement among their constituent cell-level predictions than homogeneous populations. This was quantified using generalized Jensen–Shannon dispersion, which measures divergence between individual cell-level species-probability distributions and their population mean. For 20-cell populations, median dispersion increased from 0.00 bits in pure populations to 1.32 bits in synthetic mixtures (**Figure 3E**), demonstrating substantially greater prediction heterogeneity in mixed populations.

Together with the greater recovery of AST-routing groups than exact species in balanced mixtures (Figure 2F), these analyses illustrate two complementary uses of the single-cell representation. When cell-level predictions are concordant, combining evidence across cells can substantially improve sample-level classification. When predictions are heterogeneous, retaining the underlying single-cell information can instead reveal mixed phenotypes and reduce the risk that a minority population is obscured by population averaging. Full synthetic-mixture generation, constituent-recovery definitions and generalized Jensen– Shannon dispersion analysis are provided in **Supplementary Methods Sections G.1-G.3**.

### Unseen organisms retain structured phenotypic relationships and support out-of-distribution rejection

We next examined model behaviour for five organisms belonging to species excluded entirely from model development: *Streptococcus agalactiae* (BSB), *Enterococcus faecalis* (EF), *Enterococcus faecium* (EFM), *Haemophilus inffuenzae* (HFLU) and *Listeria monocytogenes* (LMON). Because the classifier contained only the original 13 SC-MAPS species as possible outputs, unseen-organism predictions were necessarily distributed among the known classes. Nevertheless, their mean probability distributions were strongly non-uniform (**Figure 4A**). BSB showed preferential assignment towards SAUR, EFM towards ACAA and LMON towards PSMA, while HFLU distributed substantial probability between SAUR and SEPI. These assignments therefore represent similarity to learned phenotypic classes rather than valid species identification.

**Figure 4:**
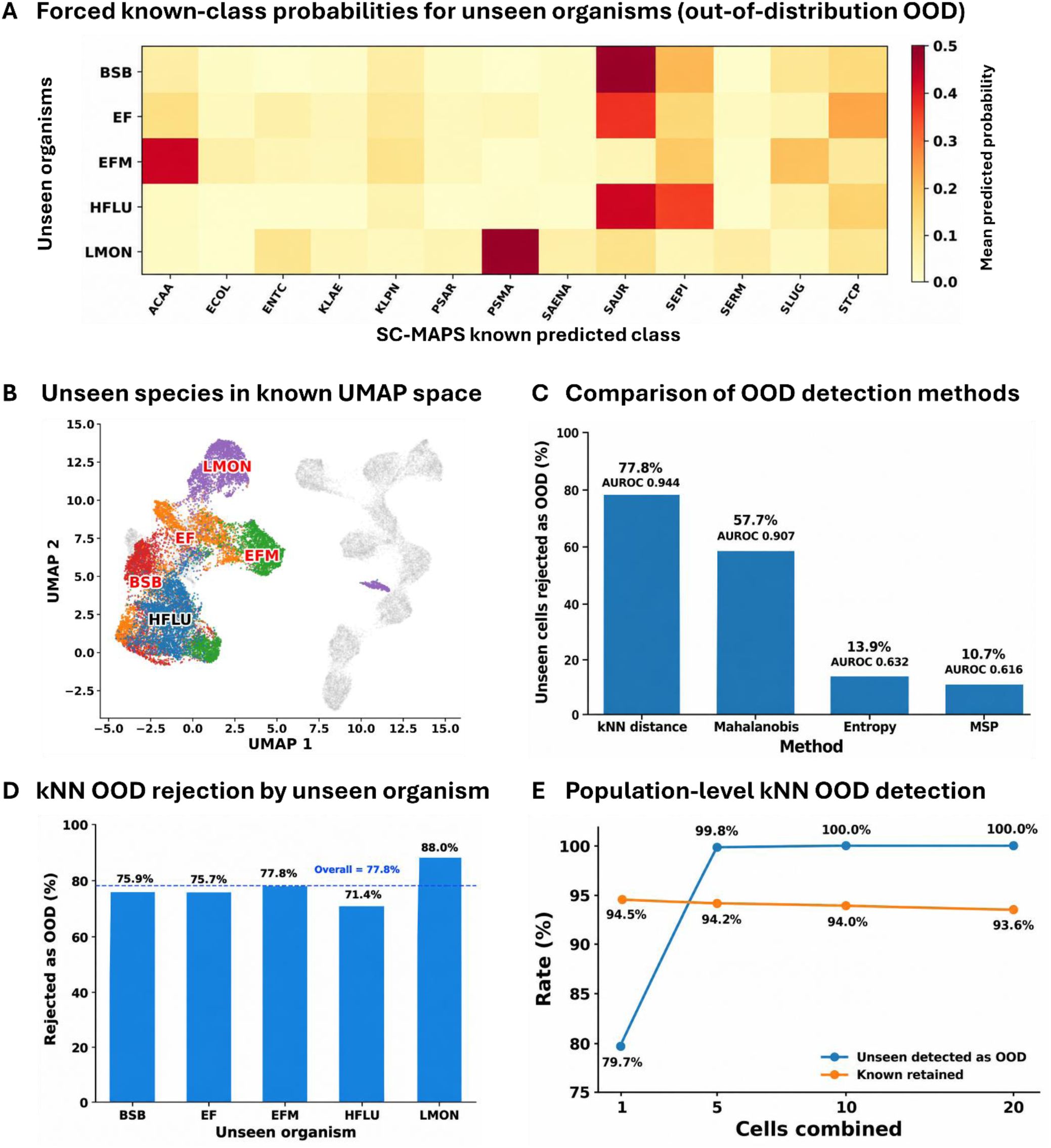
Unseen-organism behaviour and out-of-distribution detection. **(A)** Mean forced prediction probabilities for five unseen organisms across the 13 known SC-MAPS species classes. **(B)** Unseen organisms projected into the fixed known-species UMAP reference, with known cells shown in grey. **(C)** Comparison of four OOD (out-of-distribution) detection methods; kNN feature distance performed best (77.8% rejection, AUROC 0.944). **(D)** kNN OOD rejection rates for each unseen organism. **(E)** Population-level OOD detection improved from 79.7% with one cell to 99.8% with five cells and 100% with 10–20 cells, while known-cell retention remained ∼94%.

The unseen organisms were subsequently projected into the fixed known-species UMAP reference without refitting the embedding (**Figure 4B**). Their cells occupied structured regions relative to the known-species representation, consistent with their non-uniform forced probability distributions. This highlighted a limitation of closed-set classification: even an organism absent from model development must receive probability among one of the known classes.

We therefore tested whether the learned representation could support explicit out-of-distribution (OOD) rejection. Feature-space k-nearest-neighbour (kNN) distance provided the strongest single-cell discrimination between known and unseen cells, with an AUROC of 0.944 and 77.8% unseen-cell rejection at the selected operating threshold (**Figure 4C**). Mahalanobis distance achieved an AUROC of 0.907 with 57.7% rejection, whereas prediction entropy and maximum softmax probability performed substantially less well (AUROC 0.632 and 0.616, respectively). At the kNN operating point, 94.3% of known cells were retained.

OOD rejection was observed across all five unseen organisms, ranging from 71.4% for HFLU to 88.0% for LMON (**Figure 4D**). Combining evidence across cells further improved detection: unseen-organism detection increased from 79.7% in the one-cell population analysis to 99.8% with five cells and 100% with 10 or 20 cells, while known-cell retention remained approximately 94% (**Figure 4E**). These findings indicate that the learned feature representation can support an explicit unknown/OOD output rather than forcing every observation into a known species class, while phenotypic similarity to the known reference can be considered separately. Dimensionality-reduction and open-set procedures are described in **Supplementary Methods Sections 7-8**.

## Discussion

### SC-MAPS reframes bacterial classification around the downstream decision rather than perfect taxonomy

A central implication of SC-MAPS is that uncertainty in bacterial identity does not necessarily imply loss of clinically useful information. Although single-cell species classification remained challenging on strains excluded from model development, accuracy increased from 65.4% at species level to 77.1% after AST-routing group probability pooling, and 34.5% of species-level errors still mapped to the correct AST-routing group. The stronger performance at the AST-routing level suggests that broader decision-relevant phenotypic information transfers across strains more robustly than exact taxonomic discrimination. Taxonomic error and downstream decision error are therefore not equivalent: confusing organisms that share an AST pathway may be less consequential than errors between groups requiring different testing strategies. Similar principles have been reported using single-cell Raman spectroscopy, where treatment-relevant grouping can outperform exact isolate classification25–27,38. SC-MAPS extends this concept to multichannel imaging and prospectively defined AMR-informed routing groups, while not attempting to predict susceptibility, MIC or resistance mechanisms directly. Direct training on the eight routing groups did not improve performance over species-level training followed by probability pooling, suggesting that finer species supervision preserves useful phenotypic structure that can subsequently be translated into a broader clinical decision.

### Biological separability appears to matter more than increasingly complex models

The classification experiments suggest that performance was constrained more by the phenotype available to the model than by network architecture. DenseNet121, ResNet18 and ResNet34 showed broadly similar channel-dependent behaviour, whereas imaging modality had a larger effect on generalisation, and a specialist Enterobacterales classifier did not improve independent-strain holdout performance. Species separability was also uneven: ACAA, ECOL, PSAR and SLUG were comparatively well resolved, while several Enterobacterales and staphylococci remained more difficult to distinguish.

The learned DenseNet representation showed corresponding organisation when visualised using UMAP. Many species occupied recognisable regions of the embedding, while overlap remained among classes that were also difficult to separate by classification. A broader separation between Gram-positive and Gram-negative organisms was also apparent. These UMAPs were interpreted qualitatively rather than as quantitative measures of inter-species distance. Additional jointly fitted known/unseen and independently fitted unseen-only representations are provided in **Supplementary Figure S10**.

Channel-specific performance reinforced this biological interpretation. Phase contrast provided the strongest overall generalisation, while DAPI, WGA, FM1-43 and bright field contributed species-specific strengths. Multichannel fusion therefore appears to benefit from complementary biological information rather than simply increased image complexity. FM1-43 further illustrates that an informative phenotype may also be sensitive to biological or experimental variation: its strong development performance but poorer cross-strain generalisation suggests that membrane-associated fluorescence may be particularly sensitive to strain, physiological state or preparation conditions. Because strains were acquired from separate culture preparations, these effects cannot be fully disentangled here. More broadly, these findings reinforce that large cell numbers cannot substitute for biological independence. To further explore how the multichannel model uses image information, we performed an exploratory SmoothGrad–Integrated Gradients analysis on 26 correctly classified holdout cells (two per species; **Supplementary Fig. S13**). Attribution patterns varied across species and imaging channels, while phase-contrast attributions were preferentially concentrated at bacterial cell boundaries. These observations provide descriptive insights into model behaviour but do not establish causal feature importance or specific biological mechanisms.

### Multi-cell aggregation substantially improves population-level inference

The aggregation results suggest that the most useful diagnostic unit may be a small bacterial population rather than an isolated cell. A single bacterium captures only one morphological and physiological state, and may also represent only one temporal point in the cell cycle. Combining several cells instead provides repeated sampling of the population phenotype. This principle is consistent with other optical approaches, including quantitative-phase and Raman-based identification.

SC-MAPS showed the same effect using conventional multichannel microscopy: AST-routing accuracy exceeded 90% with five cells and 96% with twenty. These gains arose from combining probability distributions from individual cells rather than retraining the classifier. The value of single-cell imaging may therefore lie less in perfectly classifying every bacterium and more in preserving cell-to-cell variation before evidence is combined. This permits robust aggregation when populations are homogeneous while retaining the ability to detect heterogeneity when they are not.

### Unseen organisms reveal transferable phenotype structure and support open-set rejection

The unseen-organism analysis tested whether the learned representation contained useful structure beyond the 13 species used for model development. Cells from all five unseen organisms showed non-uniform prediction distributions and occupied characteristic regions when projected into the fixed known-species UMAP reference. Because the classifier is closed-set, however, these assignments represent similarity to learned phenotypes rather than successful identification.

Importantly, the underlying feature representation also contained information that could be used to recognise organisms outside the known reference. k-nearest-neighbour distance calculated from PCA-reduced DenseNet feature representations distinguished known from unseen cells with an AUROC of 0.944, rejecting 77.8% of unseen cells while retaining 94.3% of known cells. Population-level OOD detection increased to 99.8% when novelty scores from five cells were averaged. Feature-space distance substantially outperformed SoftMax confidence and prediction entropy, indicating that unfamiliarity was better captured by position within the learned representation than by classifier confidence alone. This is important for closed-set models, in which a confident prediction does not necessarily imply that the organism belongs to a validated class.

These findings suggest a potential framework in which samples outside the validated reference are first flagged as unknown/OOD, while their closest learned phenotypic relationship is considered separately. Such an approach could support an expanding reference atlas as additional organisms are independently characterised and incorporated into the validated class set. The present analysis included only five unseen organisms and should therefore be regarded as proof of principle rather than validation of a general open-set classifier.

### Polymicrobial analysis defines when aggregation is appropriate

The synthetic polymicrobial analysis highlights an important limitation of population averaging. Minority-species recovery was strongly abundance-dependent, whereas dominant-species recovery remained comparatively stable. In 200-cell populations, recovery of both constituent species among the two highest-ranked predictions increased from 16.3% at 5% minority abundance to 77.0% in balanced mixtures. This suggests that aggregation can reinforce the dominant population while progressively obscuring less abundant constituents. Recovery is also likely to depend on the phenotypic similarity of the organisms involved, although this relationship was not explicitly quantified here.

Retaining single-cell predictions may therefore provide a safeguard against inappropriate averaging. Mixed populations showed substantially greater disagreement among cell-level probability distributions than pure populations, with median generalized Jensen–Shannon dispersion increasing from 0.00 to 1.32 bits. This supports a framework in which prediction heterogeneity is assessed before aggregation: concordant populations can benefit from combining evidence across cells, whereas heterogeneous populations may warrant examination of their constituent single-cell predictions. AST-routing information was also more robust than exact taxonomy. In balanced mixtures containing organisms from different routing groups, both correct AST-routing groups were recovered in 95.4% of mixtures compared with 77.0% recovery of both exact species. This exceeded the 78.9% mean recovery obtained under group-size-matched randomisation (empirical *P* = 0.0015), indicating that the improvement was not explained solely by collapsing species into broader categories. These mixtures were nevertheless generated computationally from independently imaged monocultures and therefore do not capture physical co-culture, staining interactions or sample-matrix effects.

### Broader strain diversity, external validation and phenotypic AST are the critical next steps

The independent-strain holdout provides evidence that SC-MAPS phenotypes can generalise beyond strains used for model development, but the number of strains represented per species remains limited. Larger collections spanning greater within-species diversity will therefore be needed to establish the breadth of cross-strain generalisation. Architecture differences were modest, the specialist Enterobacterales experiment did not improve independent-holdout performance, and some species remained difficult across modelling strategies. Future work should therefore prioritise broader isolate collections and external evaluation across operators, laboratories, culture conditions and imaging systems.

The open-set analysis adds a second requirement for clinical translation: a deployable system should not only classify organisms it recognises but also determine when a sample falls outside the validated reference. The current feature-distance analysis suggests this may be feasible within SC-MAPS, but thresholds will require validation across substantially larger collections of known and unseen organisms.

A further extension is to combine SC-MAPS with direct phenotypic AST and evaluate performance in clinically relevant sample backgrounds. Previous microscopy studies have shown that short antibiotic exposures can produce measurable morphological and intracellular changes ^16^. SC-MAPS addresses the complementary pre-treatment question of what untreated cellular phenotype can reveal about organism identity and initial AST routing. Combining these approaches could ultimately connect organism phenotype, identification, AST routing, open-set recognition and direct antimicrobial response within a single imaging workflow.

## Methods

### Species selection and definition of AST-routing groups

Thirteen bacterial species were selected to provide a clinically relevant and taxonomically diverse panel spanning major Gram-negative and Gram-positive organisms encountered in diagnostic microbiology: *Acinetobacter baumannii* (ACAA), *Escherichia coli* (ECOL), *Enterobacter cloacae* (ENTC), *Klebsiella aerogenes* (KLAE), *Klebsiella pneumoniae* (KLPN), *Pseudomonas aeruginosa* (PSAR), *Stenotrophomonas maltophilia* (PSMA), *Salmonella enterica* (SAENA), *Serratia marcescens* (SERM), *Staphylococcus aureus* (SAUR), *Staphylococcus epidermidis* (SEPI), *Staphylococcus lugdunensis* (SLUG) and *Staphylococcus capitis* (STCP). These strains are all derived from clinical isolates rather than laboratory strains.

Species were grouped *a priori* into eight clinically informed AST-routing categories according to broad similarities and differences in their initial antimicrobial susceptibility-testing requirements: routine Enterobacterales (ECOL, KLPN), AmpC/caution Enterobacterales (KLAE, ENTC, SERM), Salmonella (SAENA), Pseudomonas (PSAR), Acinetobacter (ACAA), Stenotrophomonas (PSMA), an *S. aureus*-like group (SAUR, SLUG) and non-*lugdunensis* coagulase-negative staphylococci (CoNS; SEPI, STCP). Groups were defined to combine species for which identification would lead to a broadly similar initial AST pathway while retaining separate categories where organism identity carries distinct resistance or testing considerations ^40,41^. KLAE, ENTC and SERM were separated from routine Enterobacterales because of clinically relevant chromosomal AmpC β-lactamase biology and associated interpretive considerations^42,43^. SAENA, PSAR, ACAA and PSMA were retained as separate categories because of their distinct susceptibility-testing and resistance characteristics^44–47^. SLUG was grouped with SAUR because of its comparatively invasive clinical behaviour relative to other CoNS, while SEPI and STCP formed the non-lugdunensis CoNS group^48^. These study-defined categories were used to evaluate whether image-derived species predictions retained information relevant to initial AST-routing decisions. They were not intended as a validated clinical testing algorithm and do not represent predictions of antimicrobial susceptibility, minimum inhibitory concentration (MIC), resistance mechanisms or treatment response. The complete species composition and clinical rationale for each group are provided in **Supplementary Table 2**.

### Bacterial culture and sample preparation

The bacterial strains used in SC-MAPS, together with their corresponding plate assignments and dataset partitions, are listed in Supplementary Table 1. Each plate designation represented a distinct strain of the corresponding species.

Isolates were recovered from cryogenic stocks onto Columbia agar base supplemented with 5% (v/v) defibrinated sheep blood and incubated overnight at 37 °C. For each species, up to four distinct strains were processed independently, with each strain corresponding to one plate designation (Plates 1–4). Colonies were picked from the overnight agar culture and dispersed in Mueller–Hinton broth before fixation, without an intervening outgrowth step to a defined optical density. Aliquots of 200 µL were transferred to 96-well plates, fixed with 4% formaldehyde for 20 min, pelleted and washed before fluorescent labelling (**Figure 5A**).

**Figure 5:**
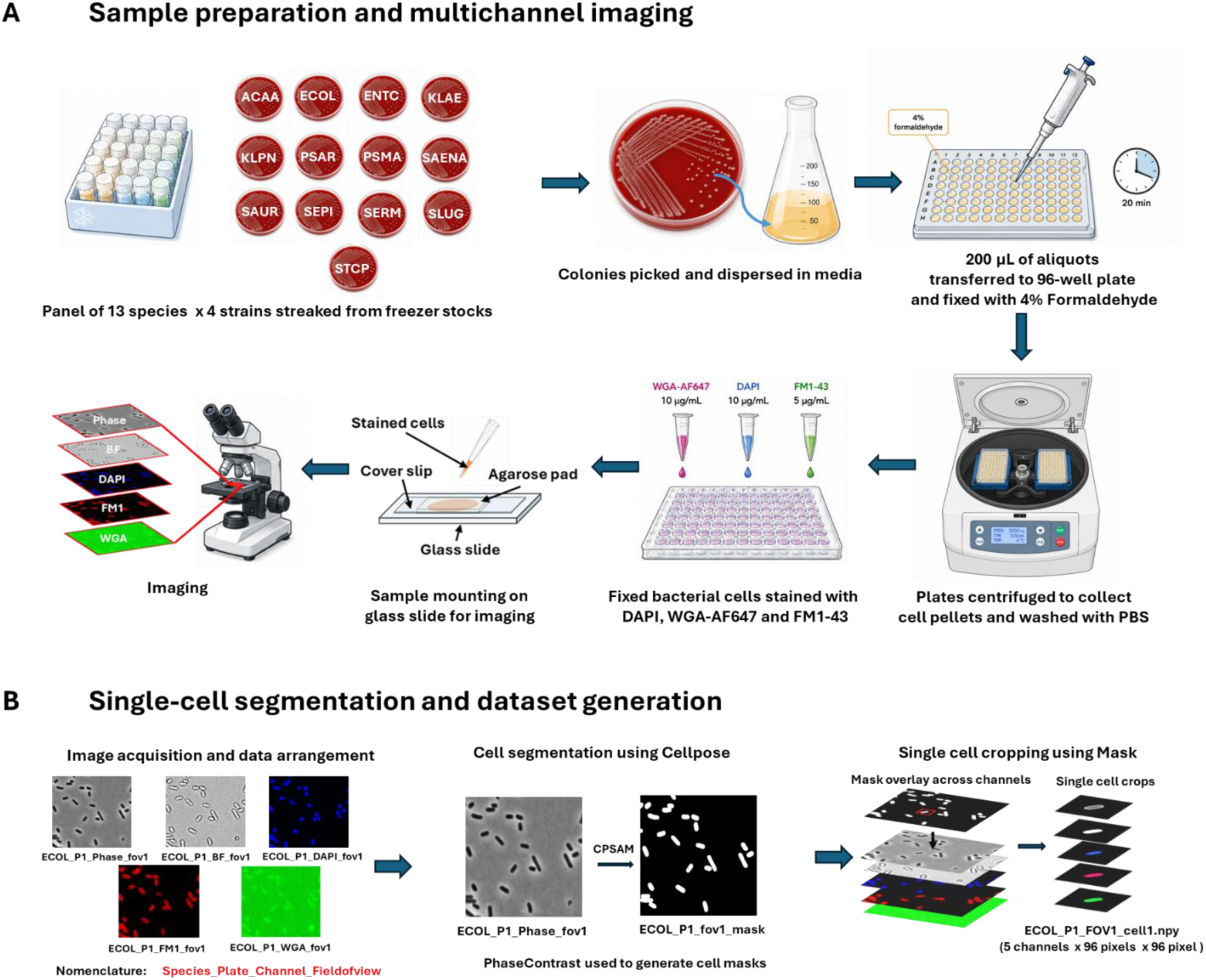
Experimental and image-processing workffow for generation of the SC-MAPS single-cell dataset. **(A)** Bacterial isolates from 13 species were recovered from cryogenic stocks on blood agar, colonies were dispersed in Mueller–Hinton broth and fixed with 4% formaldehyde for 20 min. Following pelleting and washing, cells were labelled with WGA-AF647 (10 µg/mL), DAPI (10 µg/mL) and FM1-43 (5 µg/mL), mounted on agarose pads and imaged using the Trillian microscopy system to acquire spatially matched phase-contrast, bright-field (BF), DAPI, WGA-AF647 and FM1-43 images. **(B)** Matched multichannel microscopy fields were processed at single-cell resolution. Phase-contrast images were segmented to generate instance masks, which were matched to the corresponding regions in all five imaging channels using common cell and field identifiers. The same cell mask was applied across channels to generate aligned, native-scale 96 × 96-pixel single-cell image stacks used for downstream phenotypic analysis and deep-learning classification.

Fixed bacterial cells were stained with DAPI (10 µg/mL), FM1-43 (5 µg/mL) and wheat-germ agglutinin conjugated to Alexa Fluor 647 (WGA-AF647; 10 µg/mL) to provide complementary information on intracellular and cell-envelope morphology (**Figure 5A**). DAPI is a DNA-binding fluorescent dye and was used to visualise the bacterial nucleoid, allowing differences in nucleoid size, number, spatial distribution and degree of compaction within individual cells to be captured. FM1-43 is a lipophilic styryl dye that associates with lipid membranes and was used to delineate the bacterial cell membrane, providing information on overall cell shape, membrane morphology and division-associated structures. WGA is a lectin with affinity for N-acetylglucosamine-containing structures and, when conjugated to Alexa Fluor 647, provided an additional readout of cell-wall architecture and surface-associated morphology. The relative staining patterns obtained with WGA-AF647 and FM1-43 were expected to differ according to cell-envelope composition and therefore provided complementary structural information across Gram-positive and Gram-negative organisms. Together, the DAPI, FM1-43 and WGA-AF647 fluorescence channels captured nucleoid, membrane and cell-wall-associated phenotypes, respectively, generating multichannel representations of individual bacterial cells for subsequent image-based classification.

Following staining and washing, bacterial suspensions were mounted on 1.5% (w/v) agarose pads on glass slides and covered with coverslips for microscopy. Multichannel imaging was performed using the Trillian wide-field microscopy system built around a ZEISS Axio Observer.Z1/7 inverted microscope equipped with a Plan-Apochromat 63×/1.40 NA oil-immersion Ph3 objective and an Axiocam 506m camera with a 1× camera adapter. The objective had a working distance of 193 µm and was used with immersion oil of refractive index 1.518. Images were acquired at 1 × 1 binning with a camera pixel size of 4.54 × 4.54 µm, corresponding to an effective sample-plane pixel size of approximately 0.072 µm.

Fluorescence images were acquired by wide-field epifluorescence using the 90 HE DAPI/GFP/Cy3/Cy5 reflector set. DAPI was imaged using 353-nm excitation and 465-nm emission at 30% illumination intensity, WGA-AF647 using 653-nm excitation and 668-nm emission at 30% intensity, and FM1-43 using 473-nm excitation and 579-nm emission at 20% intensity. Bright-field and phase-contrast images were acquired using transmitted white-light illumination at 10% and 30% intensity, respectively. Exposure times varied between channels and acquisitions rather than being fixed across the dataset. In representative acquisitions, exposure times were approximately 26–33 ms for DAPI, bright-field and phase contrast, 50– 100 ms for WGA-AF647, and 20 ms for FM1-43. Image acquisition was controlled using ZEN 2.6 (blue edition; version 2.6.76.00).

### Single-cell segmentation and dataset generation

Phase-contrast images were segmented using Cellpose to generate single-cell instance masks. A specialised CPSAM-based segmentation model was used for *Escherichia coli*, *Staphylococcus aureus* and *Staphylococcus epidermidis*, while the remaining species were processed using the standard Cellpose segmentation workflow. Objects intersecting image boundaries were excluded during segmentation. Images from the five imaging channels were matched using common sample and field identifiers, allowing each phase-contrast-derived instance mask to be mapped onto the corresponding bright-field (BF), DAPI, WGA-AF647 and FM1-43 images (**Figure 5B**).

Segmented objects with areas outside 40–5000 pixels were excluded to remove small fragments, segmentation artefacts and unusually large merged objects while retaining small bacterial cells. At the measured pixel size, the lower threshold corresponded to approximately 0.21 µm². These thresholds were selected following visual quality control across the species panel. For each retained cell, the segmentation-derived bounding box was expanded by 8 pixels in each direction to define the extraction region. The same instance mask was applied across all five spatially matched channels, excluding surrounding pixels and isolating the focal cell. The bounding box therefore defined the extraction region, while the segmentation mask determined which pixels were retained.

Masked single-cell crops were preserved at their native spatial resolution, centred within a 96 × 96-pixel canvas and zero-padded where necessary, without resizing. At the effective sample-plane pixel size of approximately 0.072 µm, the 96 × 96-pixel canvas corresponded to a physical area of approximately 6.9 × 6.9 µm, substantially exceeding the typical dimensions of individual bacterial cells in the dataset. This allowed single-cell crops to be accommodated at their native spatial resolution without resizing. Crops exceeding the canvas dimensions were excluded rather than rescaled. Each cell was represented by a five-channel image stack of dimensions 5 × 96 × 96 pixels, stored in the order phase contrast, BF, DAPI, WGA-AF647 and FM1-43. The image stacks were linked to a cell-level manifest containing species identity, plate/strain assignment, source image, segmentation ID and quantitative morphological and intensity measurements. Unmasked crops from the same spatial coordinates were retained separately for representative visualisation. Additional segmentation, extraction and quality-control details are provided in **Supplementary Methods Section 3** and **Supplementary Figure S1**.

The final SC-MAPS dataset comprised 916,435 segmented cells from 697 microscopy fields and 49 species–strain preparations. For each species, the strain corresponding to Plate 1 was reserved exclusively for independent holdout evaluation, while strains corresponding to Plates 2–4 were used for model development. Ten species contributed three development strains, whereas ACAA, PSMA and SEPI contributed two following image-quality exclusions. Cells originating from the same microscopy field were assigned to the same dataset partition, and no cells from the held-out strains were used during training, validation, hyperparameter optimisation or model selection. This design enabled evaluation of model generalisation to strains not encountered during development.

### Quantitative phenotypic analysis

The single-cell manifest contained morphological and imaging-derived measurements for each segmented bacterium. Species-level phenotypic profiles (**Figure 1C**) were constructed using eight morphological features—cell area, major- and minor-axis lengths, maximum Feret diameter, aspect ratio, eccentricity, solidity and compactness—together with one contrast measurement from each of the five imaging channels: phase contrast, BF, DAPI, WGA-AF647 and FM1-43. For each feature, the median was calculated across cells belonging to each species and subsequently standardised across the 13 species using z-score normalisation.

To characterise the hierarchical sources of phenotypic variability (**Figure 1D**), the same features were analysed at the single-cell level using a descriptive nested sum-of-squares variance decomposition. Total variation was partitioned into three components: between-species variation, strain/preparation-within-species variation and residual cell-within-strain/preparation variation. As each strain corresponded to a distinct culture plate and sample preparation, genetic strain-associated and preparation-associated contributions could not be independently distinguished and were therefore represented as a combined component. Each component was expressed as a percentage of the total sum of squares. This analysis was descriptive and was not intended to establish causal contributions of genetic or experimental factors to phenotypic variation. Feature definitions, standardisation procedures and variance-decomposition equations are provided in **Supplementary Methods Section 4**.

### Deep-learning classification and multichannel integration

Deep-learning analyses were performed using 96 × 96-pixel single-cell image crops. Each imaging channel was independently normalised using per-cell z-score normalisation, with the mean and standard deviation calculated across the complete 96 × 96-pixel crop, including zero-valued background and padding pixels where present. Training augmentation included horizontal and vertical flips, rotations, translations and scaling, with identical spatial transformations applied across corresponding channels to preserve image registration. Normalisation was performed after augmentation, and no augmentation was applied during validation or holdout evaluation.

DenseNet121^49^, ResNet18 and ResNet34^50^ convolutional neural networks were evaluated for 13-class species classification. Models were trained using cross-entropy loss and AdamW^51^ optimisation for up to 100 epochs. Learning-rate scheduling and early stopping were guided by validation performance, and the checkpoint achieving the highest validation macro-F1 was retained. Learning rate, weight decay, dropout and batch size were optimised using Optuna^52^, exclusively using data from the development strains. No cells from the independently held-out strains were used for training, validation, hyperparameter optimisation or checkpoint selection. The Optuna optimisation procedure, an illustrative selected configuration and additional architecture-benchmarking details are provided in **Supplementary Section 5**.

Each architecture was initially trained and evaluated independently using phase contrast, bright field (BF), DAPI, WGA-AF647 or FM1-43 as a single input channel (**Figure 6A**). Multichannel information was subsequently integrated using probability-level ensembling or feature-level fusion. For probability ensembling, species-class probabilities from independently trained single-channel models were averaged for each cell, and the species with the highest mean probability was selected (**Figure 6B**). The evaluated combinations comprised DAPI + FM1-43, DAPI + WGA-AF647 + FM1-43, DAPI + WGA-AF647 + FM1-43 + phase contrast, and all five imaging channels.

**Figure 6.**
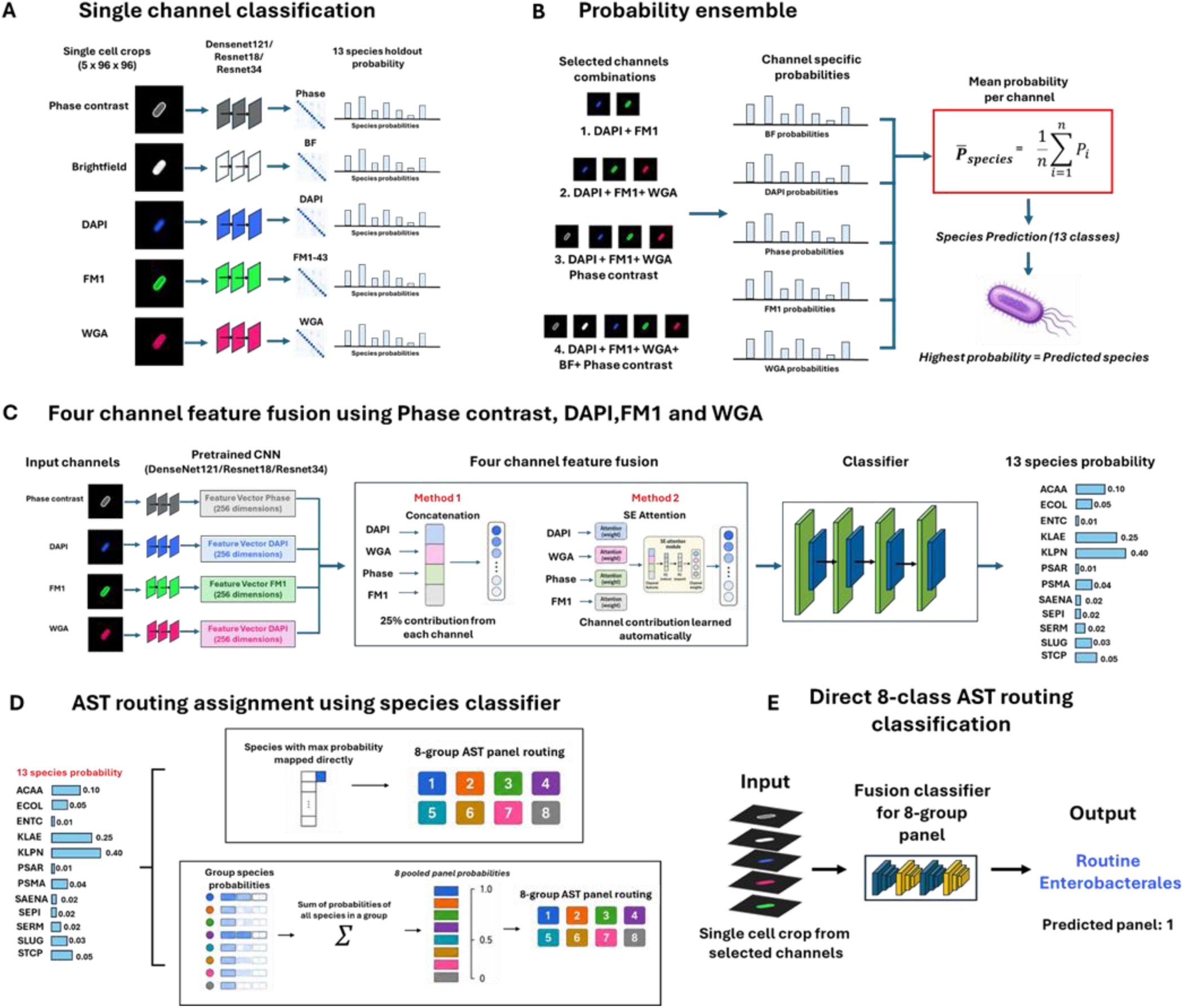
Deep-learning framework for single-channel, multichannel and AST-routing classification in SC-MAPS. **(A)** Phase-contrast, bright-field, DAPI, FM1-43 and WGA-AF647 single-cell images were analysed independently using channel-specific CNNs to generate 13-species probability distributions. **(B)** Predictions from selected combinations of independently trained channel models were combined by averaging species probabilities to generate probability-level ensemble predictions. **(C)** For feature-level fusion, features extracted from DAPI, WGA-AF647, FM1-43 and phase-contrast networks were projected to 256-dimensional representations and integrated using either direct concatenation or squeeze- and-excitation (SE) attention before 13-species classification. **(D)** Species-level outputs were converted to the eight predefined AST-routing groups either by mapping the highest-probability species to its corresponding panel or by pooling probabilities for species belonging to the same routing group. **(E)** As an alternative to species-derived routing, four-channel fusion models were trained directly to predict the eight AST-routing groups.

Feature-level fusion combined DAPI, WGA-AF647, FM1-43 and phase contrast using four channel-specific networks initialised from the corresponding previously trained single-channel classifiers (**Figure 6C**). Features from each branch were projected into 256-dimensional representations and integrated using either direct concatenation or squeeze-and-excitation (SE) attention before classification. Fusion training was performed in two stages: initially, the channel-specific branches were frozen while the fusion classification layers were trained; subsequently, higher-level feature-extraction layers were unfrozen and fine-tuned at a reduced learning rate. Both integration strategies were evaluated using DenseNet121, ResNet18 and ResNet34 architectures. The DenseNet121 concatenation model was used for subsequent AST-routing, aggregation, feature-space and unseen-organism analyses. Complete independent-strain benchmarking results for single-channel classifiers, probability ensembles and feature-fusion models are provided in **Supplementary Tables 4-6** and **Supplementary Methods Sections 5.3–5.5**.

### AST-routing classification

The predefined eight AST-routing groups were used to derive routing predictions from the 13-species classifiers using two approaches. First, the species with the highest predicted probability was assigned to its corresponding AST-routing group (top-one species mapping). Second, the full 13-species probability distribution was retained, and probabilities for species belonging to the same AST-routing group were summed. The group with the highest pooled probability was selected as the final routing prediction (**Figure 6D**). This probability-pooling approach allowed evidence distributed across multiple species within the same group to contribute to the routing decision.

To assess whether AST-routing groups could instead be predicted directly from the microscopy images, DenseNet121 feature-fusion models were trained to classify cells into the eight predefined groups rather than the 13 individual species (**Figure 6E**). These models used the same imaging data, strain-level dataset partitions and overall training framework as the species classifiers. Directly trained eight-class models were evaluated on the same independent-strain holdout and compared with routing predictions derived from the species-trained models.

To determine whether the observed routing performance could be explained by reducing the number of prediction categories, a group-size-matched randomisation analysis was performed. Species were reassigned to eight groups while preserving the number of species in each original AST-routing group. Routing performance under these randomised assignments was compared with that obtained using the clinically informed group definitions. Complete comparisons of routing strategies and randomisation procedures are provided in **Supplementary Table 7** and **Supplementary Methods Section 6**.

### Learned feature-space and unseen-organism analysis

To visualise the learned multichannel feature representation, features were extracted from the penultimate layer of the DenseNet121 concatenation model. A class-balanced subset of 2,000 independently held-out cells per species was selected, comprising 26,000 cells across the 13 known SC-MAPS species. Feature vectors were reduced to 50 dimensions using principal-component analysis (PCA) and subsequently embedded into two dimensions using Uniform Manifold Approximation and Projection (UMAP^53^). UMAP was performed using 30 nearest neighbours, a minimum distance of 0.15, Euclidean distance and a fixed random seed of 42. The embedding was fitted using the 13 known species and served as the reference representation shown in Figure 3B. Two complementary UMAP visualisations were additionally generated: a joint embedding of known and unseen organisms and an independently fitted embedding of the five unseen organisms (**Supplementary Figure S10**). The projection procedures and their limitations are described in Supplementary Section 8.

Five additional bacterial species—*Streptococcus agalactiae* (BSB), *Enterococcus faecalis* (EF), *Enterococcus faecium* (EFM), *Haemophilus inffuenzae* (HFLU) and *Listeria monocytogenes* (LMON)—were excluded from model development and subsequently processed using the same microscopy and single-cell extraction workflow. Their strain identities are provided in **Supplementary Table 8**. No retraining or model adaptation was performed. As the classifier was trained exclusively on the original 13 SC-MAPS species, predictions for unseen organisms were necessarily distributed among these known classes. These closed-set probability distributions were interpreted as model-assigneda affinities to learned species representations rather than valid identifications of the unseen organisms. Mean predicted species-probability distributions were calculated separately for each unseen organism (**Figure 4A**).

To examine the positions of unseen organisms within the learned feature space, 2,000 cells per unseen organism were transformed using the same PCA model and projected into the previously fitted known-species UMAP embedding without refitting (**Figure 4B**). This approach preserved the positions of the 13 known species, enabling the unseen organisms to be visualised relative to a common reference representation. UMAP projections were interpreted qualitatively and were not used as quantitative measures of inter-species phenotypic similarity or distance. For descriptive AST-routing analysis, the forced 13-species probability distributions of unseen organisms were pooled into the predefined AST-routing groups using the same procedure applied to the known-species holdout. These assignments were interpreted as model preferences among existing routing groups rather than clinically valid routing recommendations for organisms outside the training set. Additional details of unseen-organism cell numbers, probability analysis and feature-space projection are provided in **Supplementary Methods Sections 7-8.**

### Open-set and out-of-distribution analysis

To investigate whether organisms absent from model development could be distinguished from the 13 known SC-MAPS species, exploratory out-of-distribution (OOD) detection was performed using classifier probabilities and penultimate-layer feature representations from the DenseNet121 concatenation model. Four complementary OOD scoring methods were evaluated: maximum SoftMax probability (MSP), normalised prediction entropy, k-nearest-neighbour (kNN) feature distance and class-conditional Mahalanobis distance. For feature-based methods, representations were standardised and reduced to 50 dimensions using principal-component analysis (PCA). The kNN score was calculated as the mean Euclidean distance to the 25 nearest cells in the known-species reference set, while Mahalanobis distance was calculated relative to the known-species feature distributions. These approaches assessed prediction uncertainty or deviation from the learned feature representation rather than attempting to identify the unseen organisms.

OOD detection thresholds were determined using known-species calibration data only, without incorporating cells from the five unseen organisms during threshold selection. The primary operating point was selected to retain 95% of known calibration cells. Detection performance was evaluated using AUROC, known-species retention and unseen-organism rejection rates (**Figure 4C**). Rejection rates were additionally assessed separately for each unseen organism (**Figure 4D**).

To assess whether combining evidence from multiple cells improved OOD detection, cell-level novelty scores were averaged across populations containing 1, 5, 10, 20 or 50 cells, with 200 repeated subsamples evaluated for each population size (**Figure 4E**). Population-level detection performance was assessed alongside retention of known-species populations. Additional details of OOD scoring, threshold calibration and population-level evaluation are provided in **Supplementary Section 7**. Organism-specific rejection rates are shown in Figure 4D.

### Within-sample performance and multi-cell aggregation

Classification performance was evaluated across individual sample/image-acquisition units from the independently held-out strain of each species. A sample was defined by the combination of species, plate/strain, replicate and imaging series, with all cells assigned to a given sample originating from the same experimental and imaging unit. Within-sample classification was evaluated across all 134 independent holdout sample/image-acquisition units. Of these, 120 contained at least 200 segmented cells and were retained as a fixed cohort for the multi-cell aggregation analysis. To assess whether classification difficulty was consistent between fusion strategies, within-sample accuracies from the DenseNet121 concatenation and SE-attention models were compared using Spearman’s rank correlation at both species and AST-routing-group levels. Additional within-sample analyses are provided in **Supplementary Methods Sections 6.5-6.6**.

To investigate whether combining information from multiple cells improved classification, probability-based aggregation was performed using predictions from the independent-strain holdout. For each sample, species-probability vectors from randomly selected cells were averaged, and the species with the highest mean probability was assigned as the aggregated prediction. AST-routing predictions were obtained by summing the averaged species probabilities within each predefined routing group and selecting the group with the highest pooled probability.

Aggregation performance was evaluated using populations of 1, 2, 3, 5, 10, 20, 30, 50, 75, 100, 150 and 200 cells per sample (Figure 2D). Cells were randomly sampled without replacement within each population, and the procedure was repeated 200 times for each population size. The same 120 samples were used throughout, ensuring that each sample contributed equally to the overall evaluation regardless of its total cell count. Consequently, the one-cell condition represented a sample-balanced single-cell baseline rather than the cell-weighted accuracy calculated across the complete holdout dataset. Performance using all available cells from each sample was additionally calculated without random subsampling. Further details of probability averaging, repeated subsampling and sample-level aggregation consistency are provided in **Supplementary Section 6.6**.

### Synthetic mixed-species analysis

Synthetic two-species populations were constructed using single-cell species-probability predictions from the independently held-out Plate 1 strains, encompassing all 78 unordered pairwise combinations of the 13 SC-MAPS species. For each species pair, up to 10 source-sample pairs were evaluated. Synthetic populations contained 5, 10, 20, 50, 100 or 200 cells, with relative species abundances of 50:50, 75:25, 90:10 and 95:5. For asymmetric mixtures, each species was evaluated in turn as the dominant constituent (**Figure 7C**). Each source-sample-pair, abundance-ratio and population-size combination was independently resampled 200 times. Cells were sampled without replacement within each synthetic population, although individual holdout cells could contribute to different populations across independent repeats or experimental conditions. For each synthetic population, cell-level species-probability vectors were averaged to generate a single aggregated probability distribution. The two species with the highest aggregated probabilities were used to assess recovery of the constituent organisms. Minority-species recovery was evaluated using 200-cell populations across different abundance ratios (**Figure 3C**), while dominant-species recovery was additionally compared between 20- and 200-cell populations (**Figure 3D**). These analyses assessed the ability of the classifier to recover constituent species from computationally constructed mixtures rather than experimentally co-cultured bacterial populations. Complete mixture-generation procedures, sampling conditions and constituent-recovery definitions are provided in **Supplementary Methods Section G.1-G.3**.

**Figure 7.**
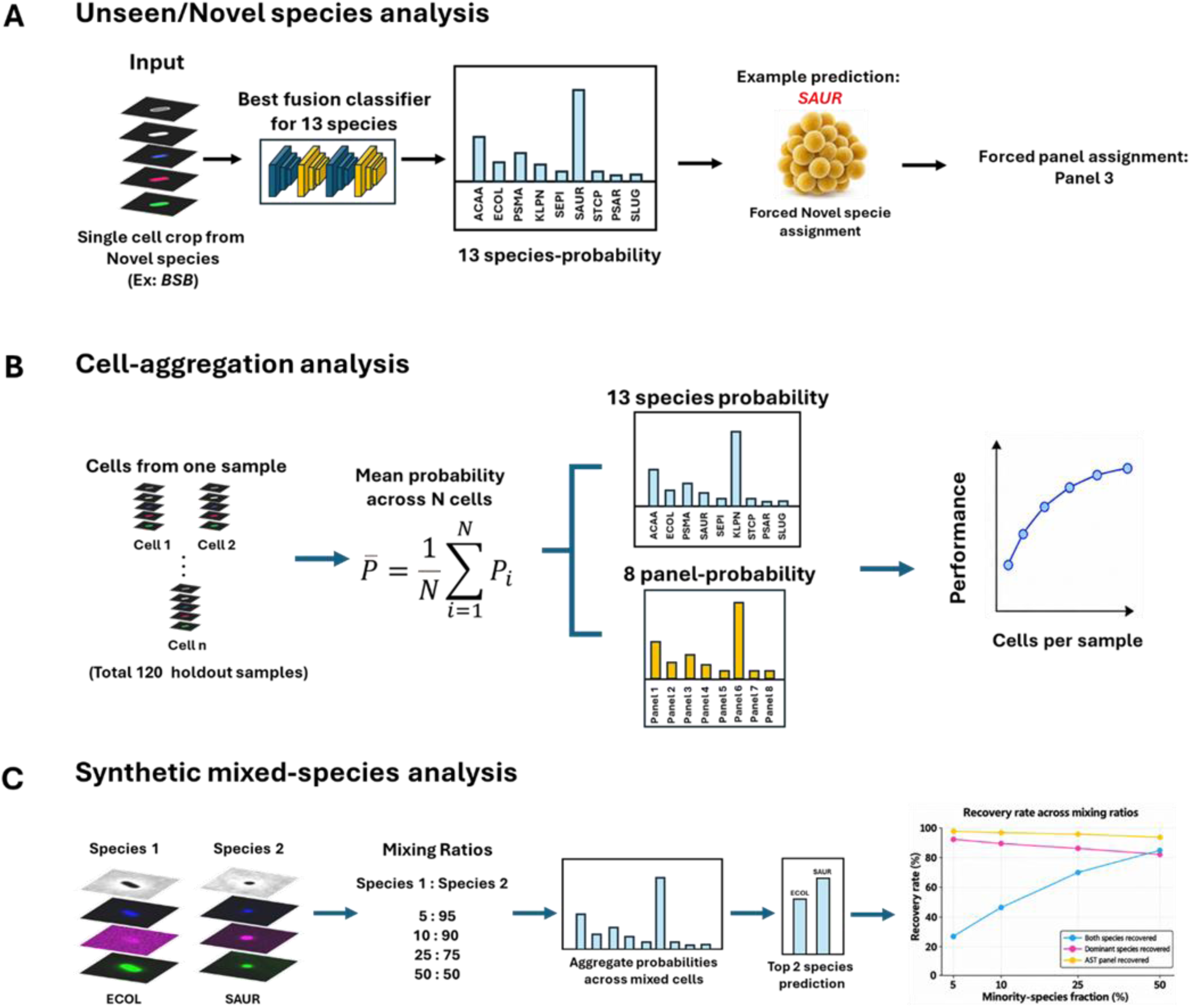
Downstream analyses used to evaluate SC-MAPS beyond single-cell species classification. **(A)** Cells from bacterial species absent from model training were processed using the trained four-channel DenseNet121 fusion model to generate forced probability distributions across the 13 known SC-MAPS species, allowing their similarity to learned species classes to be examined. **(B)** Multi-cell sample aggregation was performed by combining predicted probability distributions from increasing numbers of cells originating from the same holdout sample. Averaged species probabilities were used to generate sample-level species and AST-routing predictions, and performance was evaluated as the number of aggregated cells increased. **(C)** Synthetic mixed-species populations were generated by combining cells from the independently held-out strains of two species at defined relative abundances. Cell-level probabilities were aggregated and the two highest-probability species and corresponding AST-routing groups were evaluated to determine recovery across increasing minority-species fractions.

Prediction heterogeneity within pure and synthetic mixed populations was quantified using generalized Jensen–Shannon dispersion across the cell-level species-probability distributions. For a population containing ***N*** cells, each represented by a 13-class probability vector *p_i_*, the mean population distribution *p̅* was calculated as:

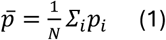

Generalized Jensen–Shannon dispersion was then defined as:

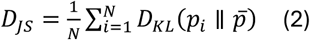

where *D_KL_* denotes the Kullback–Leibler divergence. Base-2 logarithms were used, and dispersion was therefore expressed in bits. A value of zero indicates identical probability distributions across cells, whereas larger values indicate increasing cell-to-cell disagreement. Dispersion was calculated independently for each repeated cell subsample and averaged across the 200 repeats for each source-sample or synthetic sample-pair condition. Pure and mixed populations were compared using 20-cell populations. The generalized Jensen–Shannon dispersion calculation and associated results are detailed in **Supplementary Methods Sections G.1-G.3**.

For 50:50 mixtures, recovery was additionally assessed at the AST-routing level. Species pairs were grouped according to whether the two constituent species belonged to different or the same AST-routing group. For different-routing-group pairs, recovery of both constituent species and both corresponding AST-routing groups among the two highest aggregate predictions was assessed. For same-routing-group pairs, recovery of both constituent species and of their shared AST-routing group was assessed.

### Performance metrics and statistical analysis

Classification performance was evaluated using accuracy, balanced accuracy, macro-F1, weighted F1, class-specific precision and recall, together with raw and row-normalised confusion matrices. Balanced accuracy was calculated as the mean recall across classes, while macro-F1 represented the unweighted mean of class-specific F1 scores. Macro-F1 was used as the principal model-selection metric because it gives equal weight to each class irrespective of abundance. Top-two accuracy was calculated as the proportion of predictions in which the correct species or AST-routing group was among the two highest-probability outputs.

To assess whether improvements in AST-routing accuracy exceeded those expected from reducing the number of prediction categories, group-size-matched randomisation analyses were performed. Species were randomly reassigned to eight groups while preserving the number of species in each original AST-routing group. Observed top-one species mapping, probability-pooled routing and top-two routing performance were compared with the corresponding randomised distributions (Figure 2A–C). For synthetic mixtures containing species from different routing groups, recovery of both AST-routing groups was additionally compared against 2,000 group-size-matched randomisations in which the constituent species remained assigned to different groups (**Figure 2F**). Empirical P values were determined by comparing observed performance with the corresponding randomised null distributions.

Uncertainty in AST-routing performance was assessed using sample-level bootstrap resampling of independent holdout image-acquisition units. Bootstrap-derived 95% confidence intervals were calculated for species-derived probability pooling and directly trained eight-class routing models (**Figure 2B**). For multi-cell aggregation and synthetic-mixture analyses, repeated random subsampling was used to evaluate performance across different population sizes and species compositions. Repeated subsamples from the same source population were not treated as independent biological replicates. Full randomisation, bootstrap and repeated-sampling procedures are provided in **Supplementary Methods Sections 6.3-6.4, 6.6 and G.2**.

Model training, hyperparameter optimisation, learning-rate adjustment, early stopping and checkpoint selection were performed using development-strain training and validation datasets only. For every species, all cells originating from the independently held-out Plate 1 strain were excluded from model development and reserved for final evaluation and subsequent downstream analyses.

## Data Availability

Source data underlying the main and supplementary figures will be provided with the article. The processed SC-MAPS dataset, including single-cell multichannel image crops, associated metadata and dataset-partition information are on ORA (https://dx.doi.org/10.5287/ora-nz5e6m1bw). Code used for single-cell dataset generation, deep-learning model training, evaluation, downstream analyses and figure generation will be made available at https://github.com/KapanidisLab/SC-MAPS.
All the model weights can be provided upon request. (DenseNet all channels, Resnet18 all channels, Resnet34 all channels, DenseNet121_Fusion, Resnet18_Fusion and Resnet34_Fusion)

https://github.com/KapanidisLab/SC-MAPS

https://dx.doi.org/10.5287/ora-nz5e6m1bw

## Acknowledgements

We thank Dr Maryam Beigzadeh for valuable input and discussions regarding the deep-learning analyses. We are grateful to Dr Hafez Al Sayeed for his continued scientific input and insightful discussions that helped inform the methodological approaches adopted in this study. We thank the MICRON Advanced Bioimaging Facility for access to microscopy facilities and technical support. We also acknowledge the University of Oxford Advanced Research Computing (ARC) facility for providing the high-performance computing infrastructure, GPU-accelerated computational resources and technical support used for deep-learning model training, hyperparameter optimisation and computational analyses in this study. This research used the Advanced Research Computing facilities at the University of Oxford (https://www.arc.ox.ac.uk/; https://doi.org/10.5281/zenodo.22558). This work was supported by the National Institute for Health and Care Research (NIHR) Oxford Biomedical Research Centre (BRC) (NIHR203311). The views expressed are those of the authors and not necessarily those of the NHS, the NIHR, or the Department of Health and Social Care.

## Author contributions

**A.T. and P.T. contributed equally to this work.** A.T. performed the deep-learning, computational and statistical analyses, generated the figures, interpreted the results and wrote the manuscript. P.T. generated the multichannel imaging dataset, developed the imaging workflow and contributed image-processing methods. C.F. performed the microbiological sample preparation. S.C. contributed to experimental work, methodology development and scientific guidance throughout the study. N.S., C.N. and A.N.K. contributed to study conception, supervision and interpretation of the work. N.S. contributed clinical microbiology and AST expertise, C.N. contributed machine-learning expertise, and A.N.K. provided overall project supervision and research resources. A.N.K. and N.S. contributed to funding acquisition.

All authors reviewed and edited the manuscript.

## Data and code availability

Source data underlying the main and supplementary figures will be provided with the article. The processed SC-MAPS dataset, including single-cell multichannel image crops, associated metadata and dataset-partition information are on ORA (https://dx.doi.org/10.5287/ora-nz5e6m1bw). Code used for single-cell dataset generation, deep-learning model training, evaluation, downstream analyses and figure generation will be made available at https://github.com/KapanidisLab/SC-MAPS.

All the model weights can be provided upon request. (DenseNet all channels, Resnet18 all channels, Resnet34 all channels, DenseNet121_Fusion, Resnet18_Fusion and Resnet34_Fusion)

## Supplementary Materials

All supplementary information is provided in the accompanying document SC_MAPS_SI.docx

