## Supplementary material for "SC-MAPS: Image-based deep learning for bacterial identification and antibiotic-susceptibility testing": SI File

#### 1. Bacterial species and strains information

The SC-MAPS dataset comprised 13 bacterial species, with up to four distinct strains represented for each species across four independently acquired plates. These strains are all derived from clinical isolates rather than laboratory strains. Each plate corresponded to a different strain of the respective species, providing biological variation within each species rather than repeated imaging of a single strain. Plate 1 was reserved as the independent holdout set and was not used during model development, whereas Plates 2–4 were used for training and validation. Consequently, for species represented across all four plates, model performance on Plate 1 assesses generalisation from the strains encountered during development to an independently held-out strain of the same species. The species codes and strain identities associated with each plate are provided in Supplementary Table 1.

**Supplementary Information Table 1**: Bacterial strains comprising the SC-MAPS dataset

| Species | Code | Plate 1 — Holdout strain | Plate 2 — Development strain 1 | Plate 3 — Development strain 2 | Plate 4 — Development strain 3 |
|---|---|---|---|---|---|
| *Acinetobacter baumannii* | ACAA | E10985592 | - | E30438863 | E21661965 |
| *Enterobacter cloacae* | ENTC | L121945 | E11626411 | E30770675 | E31129113_1 |
| *Escherichia coli* | ECOL | L48480 | L11037 | L64017 | L11566 |
| *Klebsiella aerogenes* | KLAE | L41994 | E11601742 | H23005962 | E31032001 |

| | | | | | |
|---|---|---|---|---|---|
| *Klebsiella pneumoniae* | KLPN | L43504 | E11635989 | E30759395 | E31199348 |
| *Pseudomonas aeruginosa* | PSAR | L46606 | E11626684 | E30778184 | E31185800 |
| *Salmonella enterica* | SAENA | L65536 | E21124276 | E30101715 | E30086536 |
| *Serratia marcescens* | SERM | L40118 | E11591769 | E21995038 | E20540962 |
| *Staphylococcus aureus* | SAUR | L44587 | E11633617 | E30766271 | E31125108_1 |
| *Staphylococcus epidermidis* | SEPI | L43925 | - | E30765807 | E31124434 |
| *Staphylococcus lugdunensis* | SLUG | L44682 | X21006231 | E30399051 | E31200012 |
| *Staphylococcus capitis* | STCP | F2015 | E11583358 | E30901604 | F23010891 |
| *Stenotrophomonas maltophilia* | PSMA | L40838 | E12061641 | - | E31147069 |

Plate 1 was used exclusively as the independent holdout set. Plates 2–4 constituted the development dataset used for model training and validation. Strain identifiers are reported as provided in the original bacterial strain records.

#### 2. AST Routing table

The 13 bacterial species were mapped into eight clinically informed antimicrobial susceptibility testing (AST) classes according to similarities in their broad downstream testing requirements. Classes were defined to group species where identification would be expected to lead to a comparable initial susceptibility-testing pathway, while retaining separate classes where organism identity is associated with substantially different resistance characteristics or antimicrobial testing considerations[1,2]. *Escherichia coli* and *Klebsiella pneumoniae* were grouped as routine Enterobacterales, whereas *Klebsiella aerogenes*, *Enterobacter cloacae* and *Serratia marcescens* formed an AmpC/caution Enterobacterales class, reflecting species-associated chromosomal AmpC β-lactamase biology[3,4]. *Salmonella enterica*, *Pseudomonas aeruginosa*, *Acinetobacter baumannii* and *Stenotrophomonas maltophilia* were maintained as separate classes because of their distinct antimicrobial susceptibility and resistance characteristics [5–8]. Among the staphylococci, *Staphylococcus aureus* and *Staphylococcus lugdunensis* were grouped into an *S. aureus*-like clinical-significance class, reflecting the unusually invasive pathogenic potential of *S. lugdunensis* relative to other coagulase-negative staphylococci, while *Staphylococcus epidermidis* and *Staphylococcus capitis* formed a CoNS (Coagulase-negative staphylococci) class[9]. These groups were used as an AMR-informed secondary classification target and were intended to represent broad AST-routing categories rather than predictions of antimicrobial susceptibility or treatment recommendations.

**Supplementary Information Table 2:** *Grouping of species together*

| AST-routing class | Species/Codes | Rationale for grouping |
|---|---|---|

| | | |
|---|---|---|
| **Routine Enterobacterales** | ECOL, KLPN | Common clinically important Enterobacterales grouped within a broad initial Enterobacterales testing pathway. Species-level identification is retained because acquired resistance mechanisms and species-specific susceptibility characteristics remain important for definitive interpretation. |
| **AmpC/caution Enterobacterales** | KLAE, ENTC, SERM | Grouped because chromosomal AmpC β-lactamase biology introduces additional β-lactam interpretive considerations. AmpC risk is species dependent, with *E. cloacae* and *K. aerogenes* showing greater propensity for clinically significant de-repression than *S. marcescens*. |
| **Salmonella-specific** | SAENA | Retained separately because *Salmonella* has organism-specific antimicrobial susceptibility considerations, particularly for detection and interpretation of fluoroquinolone resistance. |
| **Pseudomonas-specific** | PSAR | Maintained as an independent class because of its extensive intrinsic and acquired resistance mechanisms and the distinct set of antipseudomonal agents relevant to susceptibility testing. |
| **Acinetobacter-specific** | ACAA | Retained separately because of its characteristic intrinsic and acquired resistance mechanisms and antimicrobial susceptibility profile, which differs substantially from other Gram-negative non-fermenters. |
| **Stenotrophomonas-specific** | PSMA | Maintained as an independent class because of extensive intrinsic antimicrobial resistance and a comparatively restricted range of clinically relevant antimicrobial options. |
| ***S. aureus*-like clinical-significance** | SAUR, SLUG | *S. lugdunensis* is formally a coagulase-negative staphylococcus but has unusually high invasive pathogenic potential and shares important clinical characteristics with *S. aureus*, supporting its placement in an *S. aureus*-like clinical-significance group. |
| **Coagulase-negative staphylococci (CoNS)** | SEPI, STCP | Grouped as representative clinically relevant CoNS, which share a broader clinical and microbiological context distinct from *S. aureus*, particularly in device-associated and healthcare-associated infection. |

#### 3. Single cell segmentation using CPSAM

Cell segmentation was performed on phase-contrast images using Cellpose. A specialised CPSAM-based segmentation model was used for *Escherichia coli*, *Staphylococcus aureus* and *Staphylococcus epidermidis*, while the remaining species were processed using the standard Cellpose workflow. Segmentation masks were transferred to the corresponding spatially matched images across all five imaging channels using common sample, field and cell identifiers, ensuring consistent extraction of the same individual cell across modalities. Segmented objects were filtered by area, retaining cells between 40 and 5,000 pixels. Objects intersecting image boundaries were excluded. For each retained cell, the segmentation-derived bounding box was expanded by eight pixels in each direction to define the extraction region. The same binary instance mask was applied across all five channels, preserving pixel intensities within the segmented cell while setting pixels outside the mask to zero.

Masked single-cell crops were extracted at their native spatial resolution and centred within a fixed 96 × 96-pixel canvas using zero-padding. Crops exceeding these dimensions were excluded rather than resized to preserve the original image scale. Each retained cell was stored as a five-channel image stack (5 × 96 × 96 pixels), ordered as phase contrast, bright field, DAPI, WGA-AF647 and FM1-43. These image stacks were linked to a cell-level manifest containing species and strain identities, sample and acquisition identifiers, segmentation IDs, morphological measurements and channel-specific intensity features.

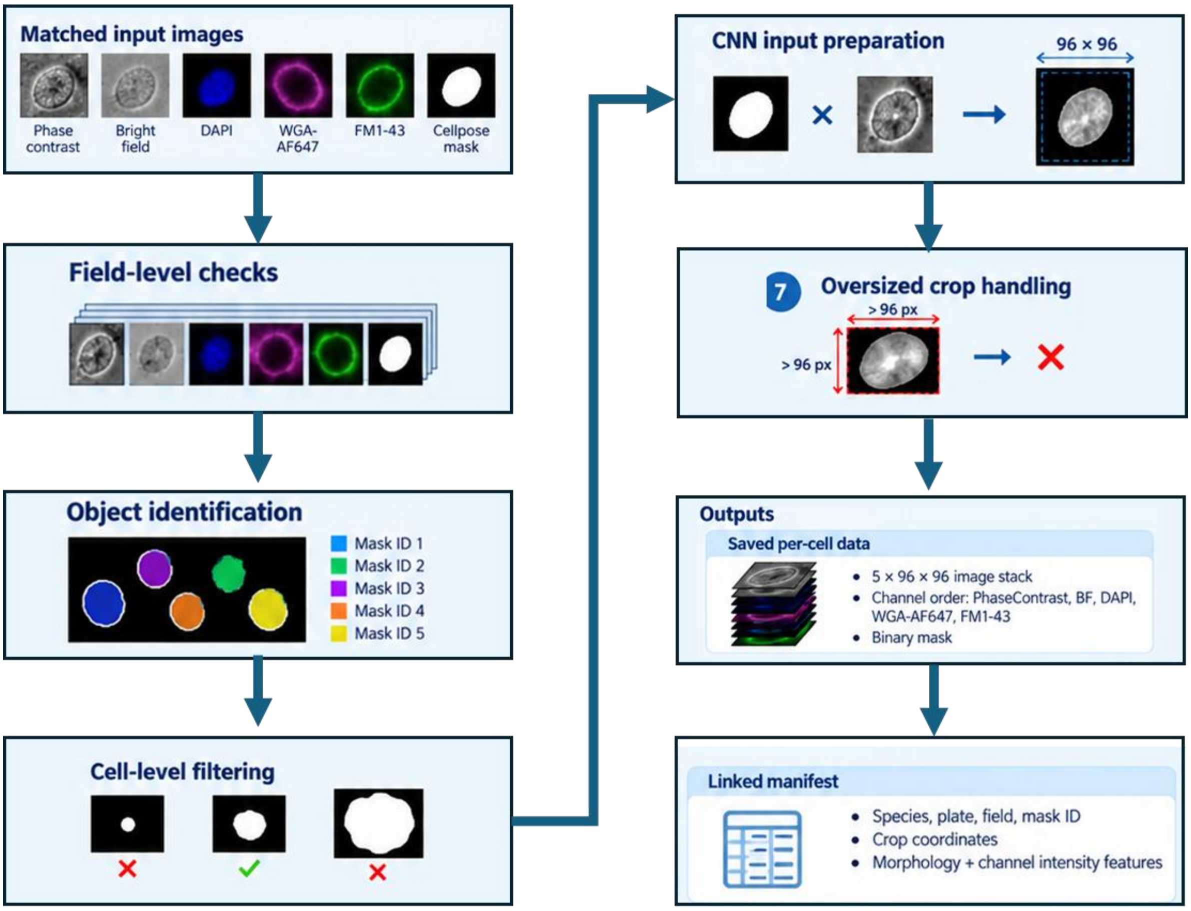

**Figure S1:** Image-processing workflow used to generate analysis-ready single-cell crops from matched multichannel microscopy images

Each extracted cell was linked to a 133-column manifest containing biological and imaging provenance, segmentation and crop-generation information, and 86 quantitative phenotypic measurements. These comprised 17 morphological descriptors, 65 channel-specific intensity and background measurements across Phase Contrast, bright field, DAPI, WGA-AF647 and FM1-43, and four cross-channel fluorescence-intensity ratios. The measurements were:

1. **Cell and sample identifiers:** unique cell ID, species, species folder, plate, dataset partition, species identifier parsed from the filename, set ID, biological replicate, technical replicate, imaging position/field, z-position, timepoint, sample key, group ID and Cellpose mask/label ID.

2. **Source-image information:** image stems and file paths for the five matched channels—Phase Contrast, bright field, DAPI, WGA-AF647 and FM1-43—as well as the associated Cellpose segmentation-mask path.

3. **Crop-generation information:** original and expanded bounding-box coordinates, native crop width and height, target canvas size, padding applied on each side, resizing flag and oversized-crop flag. These fields provided a record of how each segmented object was converted into the fixed 96 × 96 single-cell input.

4. **Morphological measurements (17 features):** *area_px, bbox_h_px, bbox_w_px, bbox_aspect_h_over_w, major_axis_length_px, minor_axis_length_px, axis_aspect_major_over_minor, eccentricity, solidity, extent, perimeter_px, orientation_rad, equivalent_diameter_px, feret_diameter_max_px, centroid_y_in_native_crop, centroid_x_in_native_crop, and compactness_4pi_area_over_perimeter2*.

5. **Channel-derived intensity measurements:** for each of the five channels, 13 measurements were calculated within the segmented cell and/or local background: *cell_mean, cell_median, cell_std, cell_min, cell_max, cell_p10, cell_p90, cell_integrated, bg_mean, bg_median, bg_std, contrast_cell_minus_bg, and contrast_cell_over_bg*. This gave 65 channel-derived features in total.

6. **Cross-channel intensity ratios (4 features):** *dapi_over_fm1_mean, wga_over_fm1_mean, dapi_over_wga_mean, and fm1_over_wga_mean,* calculated from mean signal within the segmented cell.

#### 4. Standardized morphological profiles and nested variance decomposition

Three development plates were excluded following image-quality assessment: ACAA Plate 2, PSMA Plate 3 and SEPI Plate 2, resulting in three rather than four usable strains for these species. All other species were represented by four distinct strains across the four culture plates as seen in Figure SI 2.

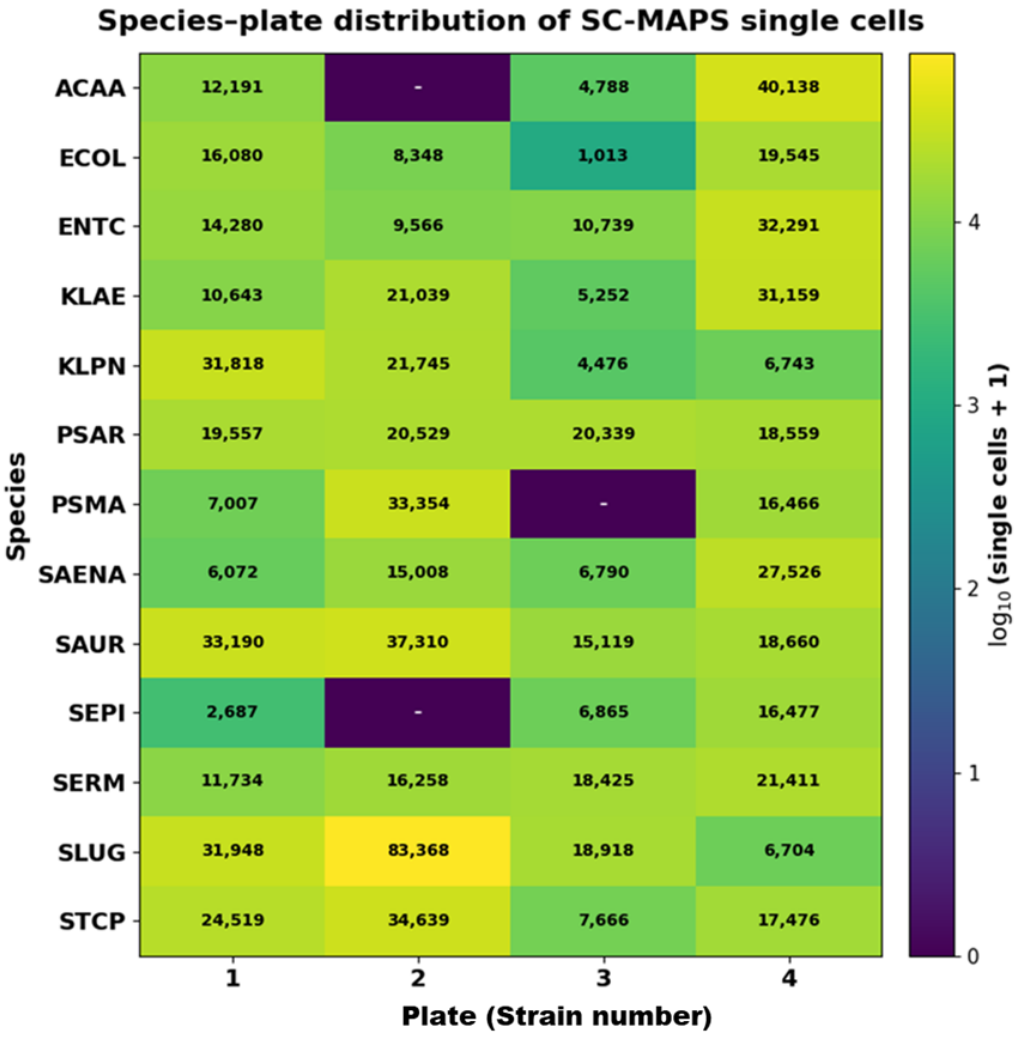

**Figure S2:** Species–strain distribution of single cells in the SC-MAPS dataset. Heatmap showing the number of retained single-cell observations for each species across four culture plates, with each plate corresponding to a distinct bacterial strain of that species

Quantitative morphological measurements were calculated from the segmentation-derived single-cell masks. For comparison of morphology across the 13 bacterial species, the median value of each feature was calculated independently for each species. The features visualised included cell area, major- and minor-axis lengths, maximum Feret diameter, aspect ratio, eccentricity, solidity and compactness. Because these measurements have different units and numerical ranges, the species-level medians were standardised independently for each feature using a z-score:

$$z_{sf} = \frac{m_{sf} - \mu_f}{\sigma_f},$$

where $m_{sf}$ is the median value of feature $f$ for species $s$, and $\mu_f$ and $\sigma_f$ are the mean and standard deviation, respectively, of the 13 species-level medians for that feature. Positive values therefore indicate a species median above the across-species mean for that feature, whereas negative values indicate a value below the across-species mean. Standardisation was performed for comparative visualisation only and was not used as input to the deep-learning models.

To describe the hierarchical structure of phenotypic variation within the dataset, a nested sum-of-squares variance decomposition was performed separately for each selected quantitative feature. Because each

culture plate corresponded to a distinct bacterial strain, variation was partitioned into components associated with species identity, strain nested within species, and residual cell-to-cell variation within each species-strain population.
For a feature value $y_{spc}$ measured for cell $c$, from strain/plate $p$ of species $s$, the total sum of squares was

$$SS_{\text{total}} = \sum_{s,p,c} \left(y_{spc} - \bar{y}\right)^2,$$

where $\bar{y}$ is the overall mean across all cells.
The species-associated component was calculated as

$$SS_{\text{species}} = \sum_{s} n_s \left(\bar{y}_s - \bar{y}\right)^2,$$

where $n_s$ is the number of cells belonging to species $s$, and $\bar{y}_s$ is the mean feature value for that species.
Variation between strains within each species was calculated as

$$SS_{\text{strain(species)}} = \sum_{s,p} n_{sp} \left(\bar{y}_{sp} - \bar{y}_s\right)^2,$$

where $n_{sp}$ is the number of cells belonging to strain $p$ within species $s$, and $\bar{y}_{sp}$ is the corresponding species-strain mean.
Residual cell-level variation was calculated as

$$SS_{\text{cell}} = \sum_{s,p,c} \left(y_{spc} - \bar{y}_{sp}\right)^2.$$

These components satisfy

$$SS_{\text{total}} = SS_{\text{species}} + SS_{\text{strain(species)}} + SS_{\text{cell}}.$$

For visualization, the contribution of each hierarchical level was expressed as a percentage of the total sum of squares:

$$P_k = 100 \times \frac{SS_k}{SS_{\text{total}}},$$

where $k$ denotes the species, strain-within-species or residual cell-level component. The resulting percentages were displayed as stacked bars in Figure 1D.

This analysis was intended as a descriptive decomposition of the observed phenotypic variation within SC-MAPS. It was not used for inferential hypothesis testing, and no $P$-values were calculated.

In addition to the eight morphological measurements, five channel-specific contrast features derived from phase contrast, bright field, DAPI, WGA-AF647 and FM1-43 were included in the species-median phenotypic profiles (Figure 1C). The same morphological and channel-contrast features were included in the nested variance-decomposition analysis (Figure 1D).

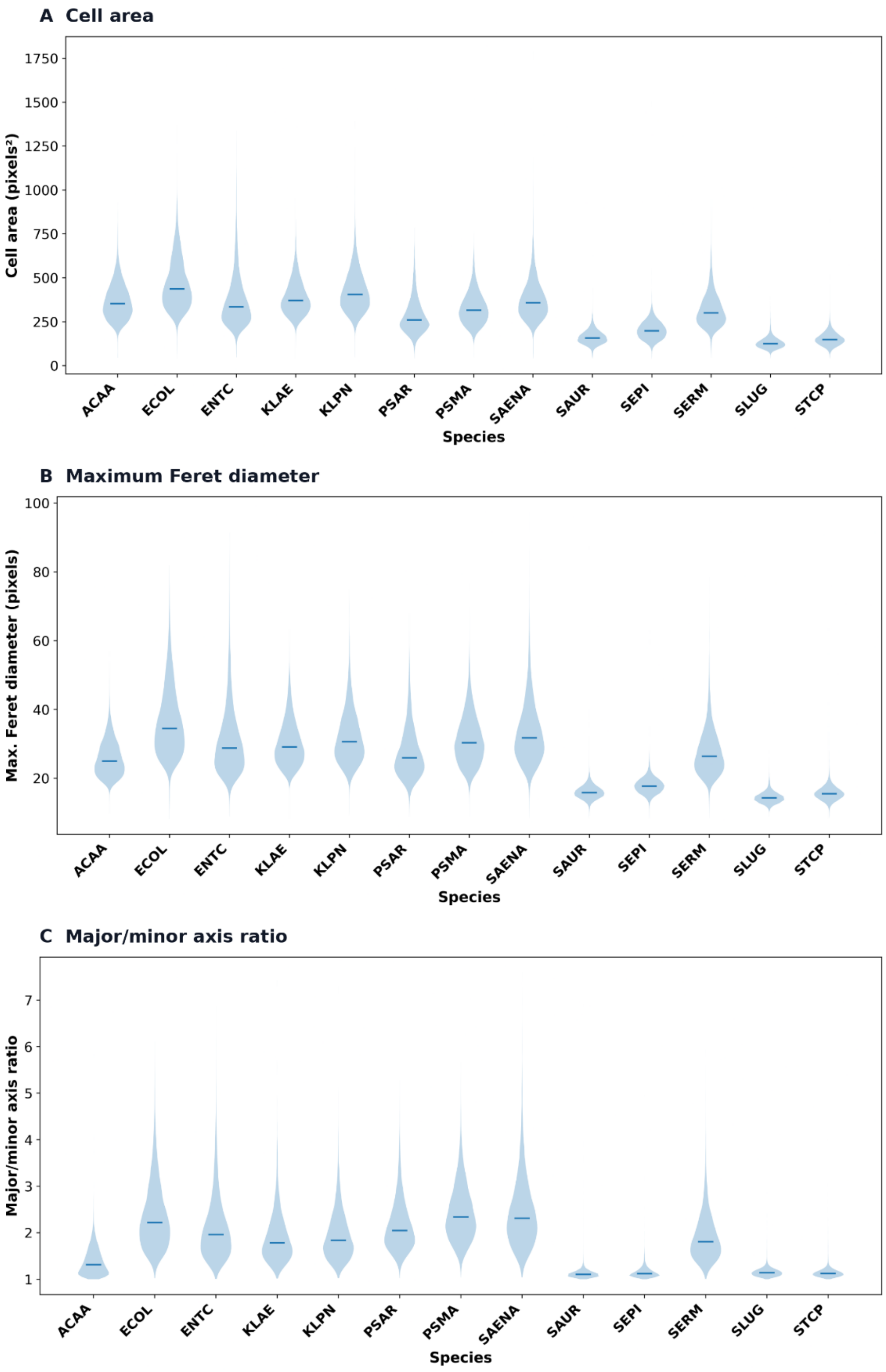

**Supplementary Figure S3: Single-cell morphological distributions across the 13 SC-MAPS bacterial species.** Violin plots showing the distributions of (A) cell area (pixels$^2$), (B) maximum Feret diameter (pixels) and (C) major-to-minor axis length ratio for individual segmented bacterial cells across the 13 species included in SC-MAPS. Violin widths represent the estimated density of single-cell measurements, while horizontal lines indicate the median for each species. The distributions illustrate species-associated differences in cell size and shape, alongside substantial morphological heterogeneity within individual species. Measurements were derived from phase-contrast segmentation masks.

#### 5. Deep-learning analysis

A systematic benchmarking framework was used to determine how imaging modality, convolutional neural network architecture and multimodal integration strategy affected bacterial classification. Benchmarking was performed progressively, beginning with single-channel models and subsequently evaluating probability-level ensembles, feature-level fusion, alternative fusion mechanisms and AST-routing classification strategies. Unless otherwise stated, all model selection was performed using the development data, while the held-out Plate 1 data were used only for independent evaluation.

The principal benchmarking stages were:

1. Single-channel classification across five imaging modalities and three CNN architectures;
2. Probability-level ensembling of complementary single-channel models;
3. Feature-level fusion of the most informative imaging channels;
4. Comparison of concatenation and squeeze-and-excitation (SE) fusion;
5. Comparison of species-derived versus directly trained AST-routing classifiers;

**Supplementary Information Table 3:** Dataset partitioning

| Partition | Number of cells | Percentage of Dataset | Biological Plate | Purpose |
|---|---|---|---|---|
| Training | 625,236 | 68.2% | 2, 3, 4 | Model fitting |
| Validation | 69,473 | 7.6% | 2, 3, 4 | Hyperparameter/model selection |
| Holdout | 221,726 | 24.2% | 1 | Independent strain level holdout |

For single-channel classification, each channel was processed independently. For multimodal feature fusion, corresponding DAPI, WGA-AF647, FM1-43 and phase-contrast crops from the same cell were provided to separate CNN branches. Bright-field imaging was evaluated during single-channel and ensemble benchmarking but was not included in the final four-channel fusion configuration because its inclusion did not improve independent-holdout performance.

Pixel values were standardised independently for each crop using z-score normalisation. Training-time image augmentation included random horizontal and vertical transformations, rotations and small spatial transformations including translation and scaling. Augmentation was applied only during model training and not during validation or holdout evaluation.

Three commonly used convolutional neural network architectures were benchmarked:

- DenseNet121[10] ;
- ResNet18[11] ; and
- ResNet34[11]

The same overall training and evaluation framework was applied across architectures to enable direct comparison of modality-dependent and architecture-dependent performance.

##### 5.1 Hyperparameter optimisation using Optuna

Hyperparameter optimisation was performed using Optuna[12] to identify suitable training configurations for the deep-learning classifiers. The search evaluated learning rate, weight decay, dropout probability, batch size and the use of class-weighted loss. For the DenseNet121 DAPI model, 12 optimisation trials were conducted, of which seven completed successfully and five were pruned before completion. The best-performing trial achieved an optimisation objective value of 0.7372, with a learning rate of $2.75 \times 10^{-4}$, weight decay of $1.16 \times 10^{-6}$, dropout probability of 0.0124 and batch size of 64, without class-weighted loss. Input images were normalised using per-crop z-score standardisation. The selected hyperparameters were subsequently used to configure model training. Hyperparameter optimisation and model selection were restricted to the development dataset, with independently held-out strains excluded from these procedures.

##### 5.2 Performance metrics and statistical analysis

Classification performance was evaluated using accuracy, balanced accuracy, precision, recall, macro-averaged F1 score (macro-F1), weighted F1 score and top-2 accuracy, as appropriate. **Accuracy** was defined as the proportion of observations assigned to the correct class. **Balanced accuracy** was calculated as the mean recall across classes, giving equal contribution to each class irrespective of class abundance. For each class, **precision** represents the proportion of predictions assigned to that class that were correct, whereas **recall** represents the proportion of observations belonging to that class that were correctly identified. The F1 score was calculated as the harmonic mean of precision and recall:

$$F1 = 2\frac{precision\ x\ recall}{\text{precision} + \text{recall}}$$

**Macro-F1** was calculated as the unweighted mean of the class-specific F1 scores and was used as the principal metric for comparing performance across classes. **Weighted F1** was calculated by weighting each class-specific F1 score according to its number of observations. **Top-2 accuracy** was defined as the proportion of observations for which the correct class occurred among the two highest predicted probabilities.

Confusion matrices were reported as raw counts or row-normalised percentages, with row-normalised values representing the proportion of observations from each true class assigned to each predicted class. For repeated multi-cell aggregation analyses, performance metrics were calculated independently for each of the 200 random subsampling repeats and subsequently summarised across repeats. Within-sample agreement between the DenseNet121 concatenation and SE-attention models was assessed using Spearman rank correlation, as described in the corresponding analysis section.

##### 5.3 Single-channel species classification

The discriminative information provided by each microscopy modality was first evaluated independently. Separate DenseNet121, ResNet18 and ResNet34 models were trained using **phase contrast, bright field, DAPI, WGA-AF647 or FM1-43** as the sole input. Identical training, validation and holdout partitions were used for every architecture–channel combination.

The resulting 15 single-channel models served two purposes. First, they enabled a controlled assessment of how strongly bacterial classification depended on imaging modality relative to CNN architecture. Second, the trained modality-specific networks provided the channel-specific models subsequently used for probability ensembling and feature-level fusion.

**Supplementary Information table 4:** Single-cell classification accuracy on individual channels

| Architecture | Channel | Holdout accuracy | Holdout macro-F1 |
|---|---|---|---|
| DenseNet121 | BF | 38.7% | 0.374 |

| | | | |
|---|---|---|---|
| DenseNet121 | DAPI | 50.9% | 0.463 |
| DenseNet121 | FM1-43 | 46.4% | 0.422 |
| DenseNet121 | Phase contrast | 54.8% | 0.488 |
| DenseNet121 | WGA-AF647 | 46.7% | 0.392 |
| ResNet18 | BF | 38.0% | 0.366 |
| ResNet18 | DAPI | 49.7% | 0.448 |
| ResNet18 | FM1-43 | 45.1% | 0.404 |
| ResNet18 | Phase contrast | 54.1% | 0.479 |
| ResNet18 | WGA-AF647 | 45.6% | 0.381 |
| ResNet34 | BF | 38.3% | 0.368 |
| ResNet34 | DAPI | 49.9% | 0.448 |
| ResNet34 | FM1-43 | 45.1% | 0.403 |
| ResNet34 | Phase contrast | 54.8% | 0.486 |
| ResNet34 | WGA-AF647 | 45.5% | 0.381 |

#### 5.4 Probability-level multichannel ensembles

Probability-level ensembling was used as the first approach for combining information from multiple imaging modalities. For each cell, the independently trained model for each selected imaging channel generated a probability distribution across the 13 species. For species *k*, probabilities were averaged across the *M* selected channels:

$$\overline{p_k} = \frac{1}{M}\sum_{m=1}^{M} p_{m,k}$$

Where $p_{m,k}$ represents the probability assigned to *k* species by imaging channel *M*. The species with the highest mean probability was selected as the ensemble prediction.

Because no additional model was trained during this procedure, probability ensembling provided a direct test of whether independently learned representations from the microscopy modalities contained complementary information. Four combinations were evaluated:

- DAPI + FM1-43
- DAPI + WGA-AF647 + FM1-43
- DAPI + WGA-AF647 + FM1-43 + phase contrast
- all five channels

Each combination was evaluated independently for DenseNet121, ResNet18 and ResNet34. The four-channel DAPI + WGA-AF647 + FM1-43 + phase-contrast combination provided the strongest and most consistent improvement and was therefore selected for subsequent feature-fusion experiments.

**Supplementary information table 5:** Probability ensemble studies across architectures

| Architecture | Channels | Holdout accuracy | Macro-F1 | Gain vs best single channel |
|---|---|---|---|---|
| DenseNet121 | DAPI + FM1 | 54.0% | 0.503 | −0.9 pp |
| DenseNet121 | DAPI + WGA + FM1 | 56.7% | 0.522 | +1.9 pp |
| DenseNet121 | DAPI + WGA + FM1 + Phase contrast | **61.2%** | **0.562** | **+6.4 pp** |
| DenseNet121 | All five | 60.0% | 0.555 | +5.2 pp |
| ResNet18 | DAPI + FM1 | 52.5% | 0.482 | −1.6 pp |
| ResNet18 | DAPI + WGA + FM1 | 54.8% | 0.494 | +0.7 pp |
| ResNet18 | DAPI + WGA + FM1 + Phase contrast | **59.1%** | **0.535** | **+5.0 pp** |
| ResNet18 | All five | 58.2% | 0.531 | +4.1 pp |
| ResNet34 | DAPI + FM1 | 52.4% | 0.476 | −2.4 pp |
| ResNet34 | DAPI + WGA + FM1 | 54.7% | 0.492 | −0.1 pp |
| ResNet34 | DAPI + WGA + FM1 + Phase contrast | **59.3%** | **0.536** | **+4.5 pp** |
| ResNet34 | All five | 58.4% | 0.534 | +3.6 pp |

#### 5.5 Multichannel feature fusion

Feature-level fusion was evaluated to determine whether integrating learned channel-specific representations provided additional information beyond averaging final classification probabilities.

For each cell, the DAPI, WGA-AF647, FM1-43 and phase-contrast images were passed independently through their corresponding pretrained single-channel CNN branches. Rather than retaining the final 13-class outputs, the higher-level representation produced by each network was extracted and projected into a common **256-dimensional feature representation**.

##### Concatenation fusion

For concatenation fusion, the four 256-dimensional feature vectors were concatenated:

$$\mathbf{z} = [\mathbf{z}_{DAPI}, \mathbf{z}_{WGA}, \mathbf{z}_{FM1}, \mathbf{z}_{PC}]$$

producing a combined 1024-dimensional representation before the shared classification head.

The concatenation strategy therefore retained the full channel-specific feature vectors and allowed the final classifier to learn directly from their joint representation.

##### SE-attention fusion

A second fusion architecture used squeeze-and-excitation attention. Channel-specific representations were adaptively reweighted before integration, enabling the model to emphasise feature components that contributed most strongly to classification. The reweighted representations were subsequently combined and passed to the classification head.

##### Two-stage training of feature-fusion models

Feature-fusion models were trained in two stages.

During **Stage 1**, the pretrained channel-specific CNN branches were frozen. Only the newly introduced feature-projection, fusion and classification layers were optimised. This allowed the multimodal classifier to learn how to combine the existing channel-specific representations without initially modifying the representations learned during single-channel training.

During **Stage 2**, the best-performing Stage 1 model was fine-tuned. The final feature-extraction section of each channel-specific CNN was unfrozen, whereas earlier network layers remained fixed. The unfrozen pretrained layers were updated at a lower learning rate than the newly introduced fusion layers. This enabled higher-level representations to adapt to the multimodal task while retaining lower-level features learned during single-channel classification.

Both concatenation and SE-attention fusion were evaluated using DenseNet121, ResNet18 and ResNet34, allowing three levels of modality integration to be compared systematically: single-channel classification, probability-level integration and feature-level integration.

**Supplementary information Table 6:** Summary of feature fusion results across architectures

| Architecture | Fusion method | Holdout accuracy | Macro-F1 | Gain vs best single channel | Gain vs best ensemble |
|---|---|---|---|---|---|
| DenseNet121 | Concatenation | **65.4%** | **0.607** | +10.5 pp | +4.2 pp |
| DenseNet121 | SE attention | 64.3% | 0.600 | +9.5 pp | +3.1 pp |
| ResNet18 | Concatenation | 61.4% | 0.574 | +7.3 pp | +2.3 pp |
| ResNet18 | SE attention | 62.1% | 0.581 | +8.0 pp | +3.0 pp |
| ResNet34 | Concatenation | 62.4% | 0.571 | +7.6 pp | +3.1 pp |
| ResNet34 | SE attention | 61.6% | 0.559 | +6.8 pp | +2.3 pp |

DenseNet121 concatenation was taken forward as the principal SC-MAPS model because it produced the strongest independent-holdout species classification.

#### 5.6 Specialist classification of difficult Enterobacterales

The multichannel fusion models showed persistent confusion among four closely related Enterobacterales classes: *Enterobacter cloacae* (ENTC), *Klebsiella aerogenes* (KLAE), *Klebsiella pneumoniae* (KLPN) and *Serratia marcescens* (SERM). We therefore tested whether restricting classification to these difficult classes could improve discrimination beyond the original 13-class fusion model.

A specialist four-class classifier was trained using the fused representations of ENTC, KLAE, KLPN and SERM, with the pretrained fusion feature extractor frozen and only the specialist classification head trained. The specialist model showed strong performance on the development data, indicating that the four classes could be separated within the strains used for model development. However, this improvement did not transfer to the independently held-out Plate 1 strains. On the corresponding four-species holdout subset, the specialist classifier achieved **51.5% accuracy and a macro-F1 score of 0.500**, and did not provide an improvement sufficient to justify replacing the predictions of the original fusion model. The specialist classifier was therefore not used in subsequent analyses.

This result suggests that the residual confusion among these Enterobacterales was not resolved simply by restricting the classification problem to a smaller number of related species. In particular, the contrast between development and independent-strain performance indicates that apparent separability within the development strains did not necessarily translate to generalisation across strains.

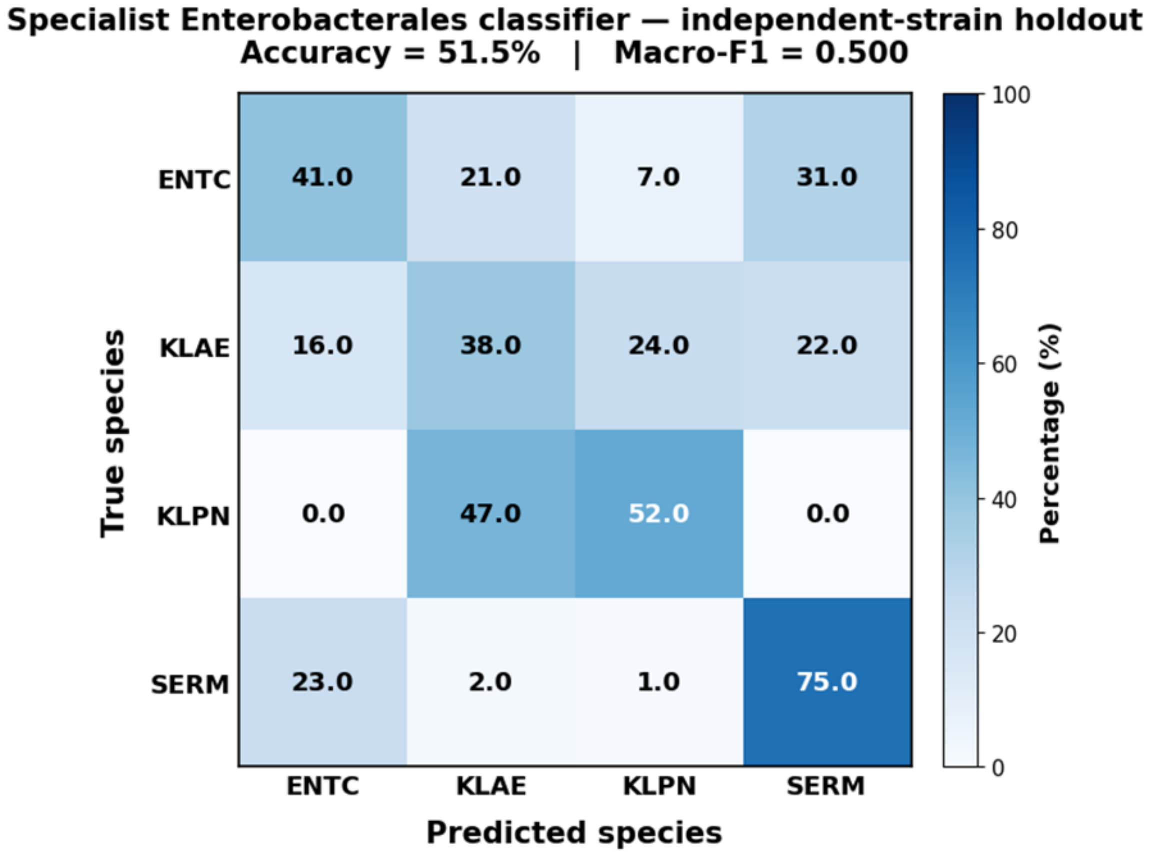

**Figure S4. Specialist Enterobacterales classification on the independent-strain holdout**. Row-normalised confusion matrix for the four-class specialist classifier (ENTC, KLAE, KLPN and SERM). Despite strong development performance, holdout accuracy was 51.5% (macro-F1 = 0.500), with substantial confusion among ENTC, KLAE and KLPN.

#### 6. AST-routing predictions

##### 6.1 AST-routing predictions derived from species classifiers

AST-routing classification was evaluated using the 13-species DenseNet121 fusion classifiers in two ways.

###### Top-1 species mapping

The species with the largest predicted probability was first selected:

$$\hat{s} = \arg\max p_s$$

and its predefined AST-routing group was then assigned. This analysis allowed species-level errors that remained within the correct routing group to be identified.

##### Probability pooling

Rather than first selecting a species, the complete 13-class probability distribution was retained. For routing group $g$, probabilities for all species assigned to that group were summed:

$$\boldsymbol{P}(\boldsymbol{g}) = \sum_{s \in g} \boldsymbol{p}_s$$

and the routing group with the highest pooled probability was selected. Probability pooling allows uncertainty distributed across multiple species belonging to the same routing group to contribute collectively to the correct downstream prediction.

#### 6.2 Direct eight-class AST-routing classification

To investigate whether direct prediction of AST-routing groups offered an advantage over species-derived routing, DenseNet121 feature-fusion models were trained to classify individual cells directly into the eight predefined AST-routing groups rather than the original 13 species. The same four imaging channels (phase contrast, DAPI, WGA-AF647 and FM1-43), strain-level dataset partitions and overall training framework were used.

The direct eight-class classifiers generated a probability distribution across the eight routing groups, with the highest-probability group selected as the final prediction. Performance was evaluated using the independent-strain holdout and compared with the top-1 mapping and probability-pooling approaches derived from the 13-species classifiers. This comparison assessed whether learning the broader routing categories directly improved classification or whether retaining species-level supervision provided a more informative representation for downstream AST routing.

**Supplementary information table 7:** AST routing results summary

| Training strategy | Prediction strategy | Holdout accuracy | Macro-F1 |
|---|---|---|---|
| 13-species DenseNet concat | Top-1 species → routing group | 76.9% | 0.746 |
| 13-species DenseNet concat | Probability pooling | **77.1%** | **0.748** |
| Direct 8-class DenseNet concat | Direct routing prediction | 74.9% | 0.737 |
| Direct 8-class DenseNet SE | Direct routing prediction | 75.0% | 0.739 |

#### 6.3 Comparison of AST-routing strategies and bootstrap uncertainty

Uncertainty in AST-routing accuracy was assessed using bootstrap resampling of independent holdout sample/image-acquisition units, preserving the clustering of cells within each sample. Classification accuracy was recalculated for each bootstrap sample, and 95% confidence intervals (CIs) were obtained from the resulting distributions. Species-derived probability pooling achieved 77.1% accuracy (95% CI: 75.5–78.8%), compared with 75.0% (95% CI: 73.0–76.9%) for direct eight-class classification (Supplementary Figure S5). These intervals describe uncertainty in the individual estimates and do not independently establish a statistically significant difference between strategies.

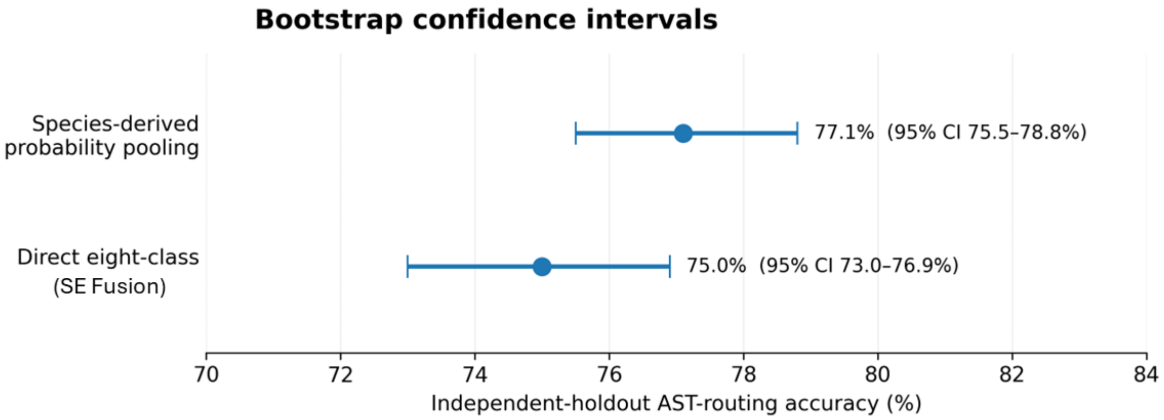

**Supplementary Figure S5: Bootstrap uncertainty in AST-routing classification performance.** Comparison of independent-strain holdout accuracy between species-derived probability pooling and directly trained eight-class AST-routing classification. Points indicate observed classification accuracies, and horizontal error bars represent bootstrap-derived 95% confidence intervals. Bootstrap resampling was performed at the sample/image-acquisition level to account for clustering of cells within experimental units. Species-derived probability pooling achieved 77.1% accuracy (95% CI: 75.5–78.8%), compared with approximately 75.0% (95% CI: 73.0–76.9%) for direct eight-class classification for SE fusion.

##### 6.4 Group-size-matched randomisation analysis

To determine whether improved AST-routing accuracy resulted simply from reducing 13 species to eight categories, species were randomly reassigned to routing groups while preserving the original group sizes. Observed top-1 species mapping, probability pooling and top-2 routing accuracies were compared with the corresponding randomised null distributions. Observed performance exceeded randomised means by 8.9, 9.0 and 10.5 percentage points, respectively (empirical $P$ = 0.0087, 0.0115 and 0.0005; Supplementary Figure S6). This analysis assessed whether the predefined AST-routing groups retained predictive information beyond that expected from category reduction alone.

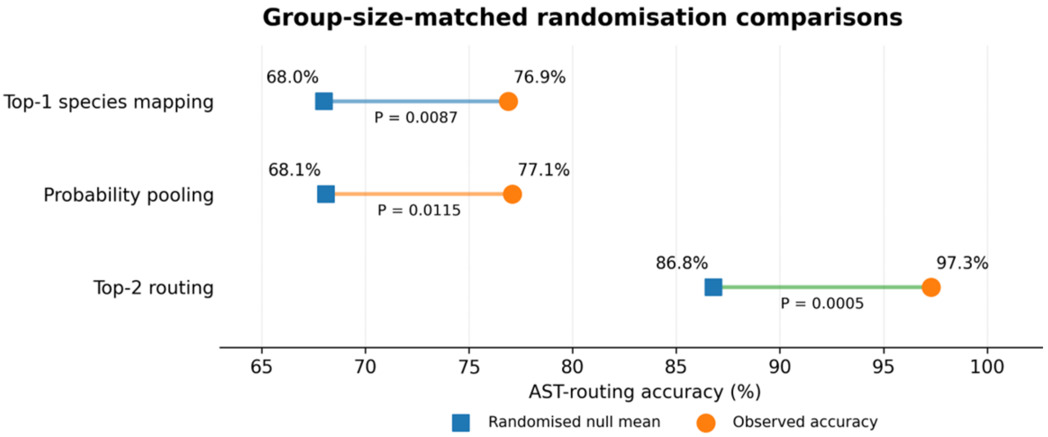

**Supplementary Figure S6: Group-size-matched randomisation analysis of AST-routing classification.** Comparison of observed AST-routing accuracy with mean accuracy obtained under randomised species-to-group assignments for top-1 species mapping, probability pooling and top-2 routing. Randomised assignments preserved the original number of species within each of the eight AST-routing groups. Observed accuracies exceeded the corresponding randomised means by 8.9, 9.0 and 10.5 percentage points, respectively (empirical $P$ = 0.0087, 0.0115 and 0.0005). The analysis assessed whether the observed routing performance exceeded that expected from grouping 13 species into eight categories alone.

##### 6.5 Within-sample classification and model consistency

Within-sample classification performance was assessed using predictions from the independently held-out Plate 1 strains. A holdout sample was defined as an individual image-acquisition unit identified by the combination of species, plate/strain, replicate and imaging series, with all constituent cells originating from the same experimental and imaging unit. The analysis included 134 holdout samples. Of these, 120 contained at least 200 segmented cells and were selected for the subsequent multi-cell aggregation analysis.

For each sample, species-level accuracy was calculated as the proportion of cells correctly assigned to their true species. AST-routing accuracy was calculated similarly, following probability pooling of the 13 species predictions into the eight predefined routing groups. This produced a separate species and AST-routing accuracy for each sample, allowing classification difficulty to be assessed independently of differences in sample size.

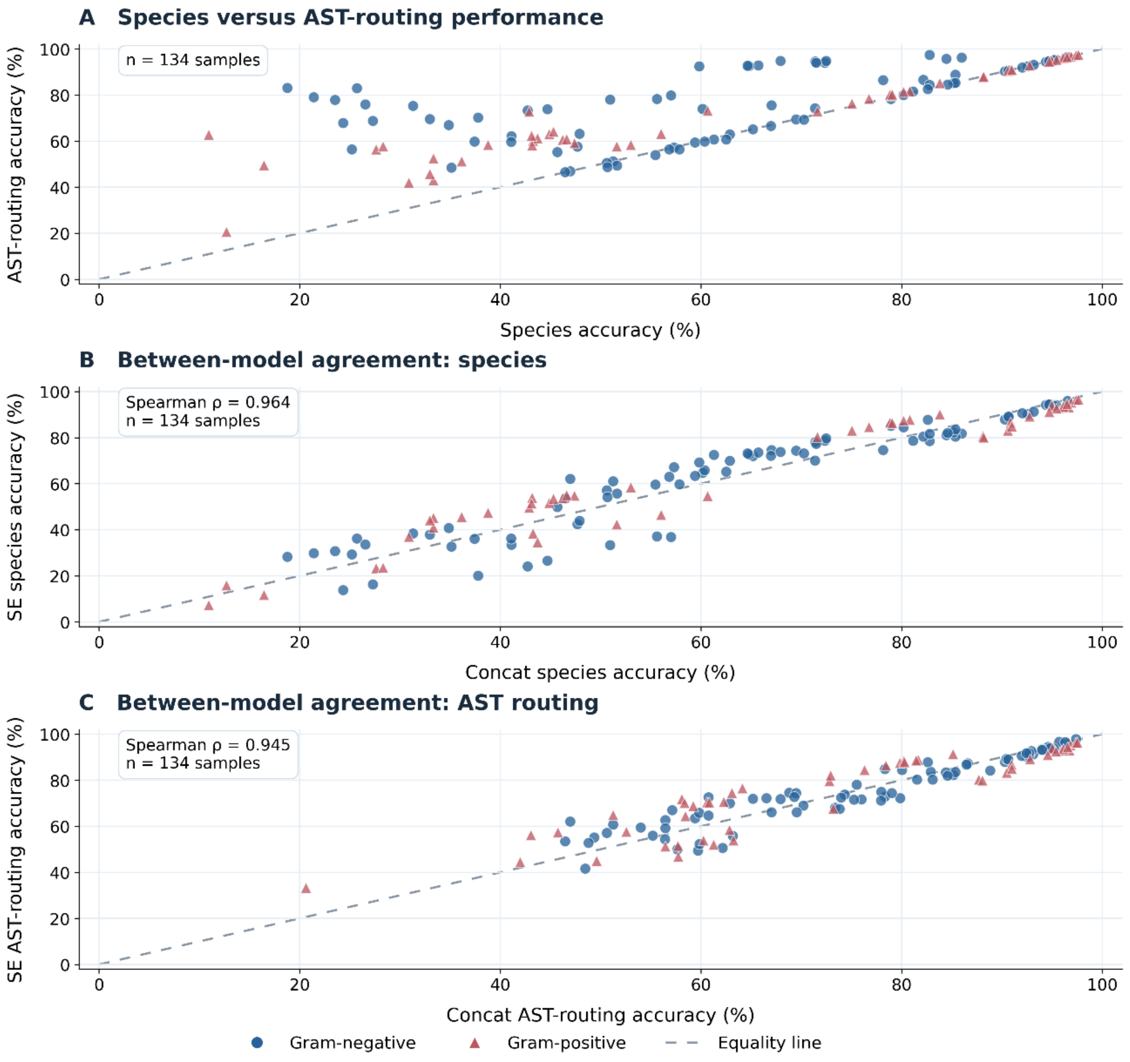

**Supplementary Figure S7: Within-sample classification performance and consistency between multichannel fusion models. (A)** Comparison of species-level and probability-pooled AST-routing accuracy for the DenseNet121 concatenation model across 134 independent holdout sample/image-acquisition units. **(B, C)** Agreement in within-sample classification accuracy between DenseNet121 concatenation and squeeze-and-excitation (SE) attention fusion models at (B) species and (C) AST-routing-group levels. Each point represents one holdout sample, with Gram-negative species shown as blue circles and Gram-positive species as red triangles. Dashed diagonal lines indicate equal accuracy between the compared measures. Within-sample performance was strongly correlated between fusion strategies at both species level (Spearman ρ = 0.964) and AST-routing level (ρ = 0.945), indicating consistent patterns of sample-level classification difficulty across the two models.

#### 6.6 Multi-cell probability aggregation

Multi-cell aggregation was performed using fixed cell-level predictions from the DenseNet121 concatenation and squeeze-and-excitation (SE) attention fusion models. For each holdout sample, predicted species-probability distributions from randomly selected cells were averaged to generate a single population-level prediction. The species with the highest mean probability was selected as the aggregated species prediction. AST-routing predictions were obtained by summing the averaged probabilities of species belonging to each predefined routing group and selecting the group with the highest pooled probability. This approach combined the complete probability distributions rather than applying majority voting to individual cell classifications.

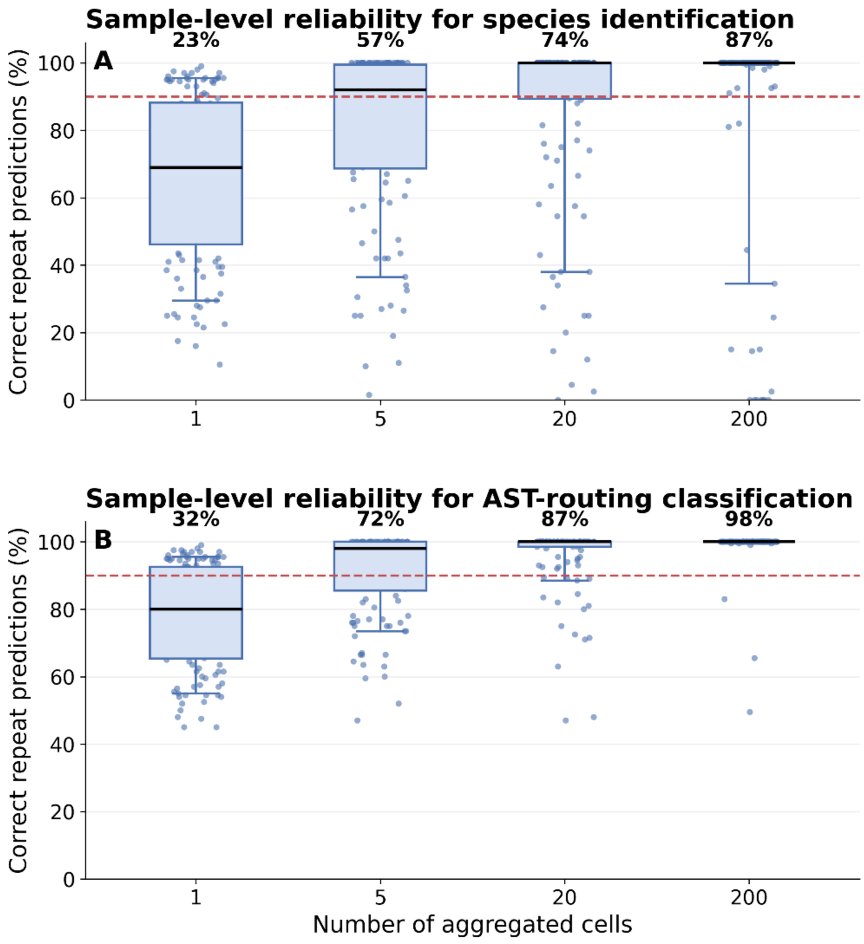

**Supplementary Figure S8: Sample-level classification consistency during multi-cell aggregation.** Distributions of correct prediction rates across 200 repeated cell sub-samplings for (A) species identification and (B) AST-routing classification using the four-channel DenseNet121 concatenation model. Each point represents one of 120 independently held-out sample/image-acquisition units, evaluated using populations of 1, 5, 20 and 200 cells. Boxes indicate medians and interquartile ranges, with whiskers extending to the 10th and 90th percentiles. The dashed red line indicates 90% correct predictions across repeats, while percentages above each distribution indicate the proportion of samples meeting or exceeding this threshold. The distributions illustrate differences in aggregation consistency between samples and the greater stability of AST-routing predictions compared with exact species identification.

Aggregation was evaluated using populations of *1*, 2, 3, 5, 10, 20, 30, 50, 75, 100, 150 and 200 cells. At each population size, cells were randomly sampled without replacement from the same 120 independently held-out samples, each containing at least 200 segmented cells. The procedure was repeated 200 times, and classification accuracy was calculated across samples for each repeat before averaging across repeats. Each sample contributed equally to the analysis regardless of its total cell count. Consequently, the one-cell aggregation baseline represented sample-balanced rather than cell-weighted accuracy. A separate deterministic analysis was performed using all available cells within each sample, without random subsampling.

**Sample-level aggregation consistency.** To investigate whether improvements in overall aggregation accuracy were consistent across individual samples, the proportion of correct predictions across the 200 subsampling repeats was calculated separately for each sample and population size (Supplementary Figure S8). At 20 aggregated cells, 89 of 120 samples (74.2%) achieved correct species identification in at least 90% of repeats, compared with 104 samples (86.7%) for AST-routing classification. At 200 cells, these proportions increased to 104 samples (86.7%) and 117 samples (97.5%), respectively. The distributions revealed considerable variation in aggregation performance between samples, with some achieving consistently correct predictions using relatively few cells while others remained difficult despite increasing population size. AST-routing predictions generally showed greater consistency than exact species classification. The 90% threshold was used as a descriptive measure of subsampling consistency rather than a clinically validated reliability threshold. Repeated subsamples from the same acquisition were not considered independent biological replicates.

#### 7. Out-of-distribution detection

To assess whether bacterial organisms excluded from model development could be distinguished from the 13 known SC-MAPS species, out-of-distribution (OOD) detection was performed using the four-channel DenseNet121 concatenation model. Five unseen organisms were evaluated: *Streptococcus agalactiae* (BSB), *Enterococcus faecalis* (EF), *Enterococcus faecium* (EFM), *Haemophilus influenzae* (HFLU) and *Listeria monocytogenes* (LMON). These organisms were excluded from model training, validation, hyperparameter optimisation and threshold calibration. OOD detection was performed using the existing trained model without retraining or adaptation.

Following single-cell extraction and quality filtering, the available populations comprised approximately 26,200 BSB, 27,500 EF, 9,500 EFM, 16,800 LMON and 4,400 HFLU cells. Their strain identities are provided in Supplementary Table 8.

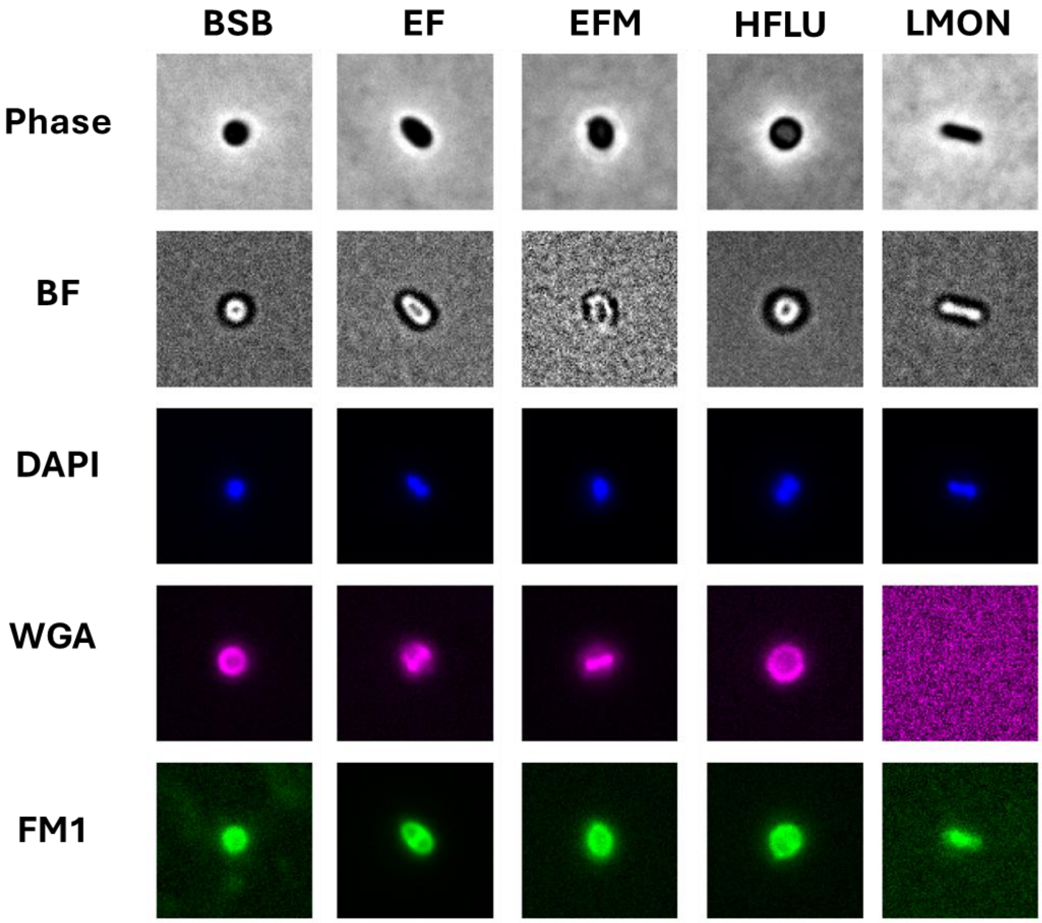

**Figure S9: Representative multichannel microscopy images of individual bacterial cells.** Spatially matched single-cell crops showing five representative bacteria imaged using phase contrast, bright field (BF), DAPI, WGA-AF647 and FM1-43 (top to bottom). Each column represents the same individual bacterium across all five imaging channels. Phase-contrast and BF images provide information on cellular morphology, while DAPI (blue) highlights nucleoid organisation, WGA-AF647 (magenta) labels cell-envelope-associated glycoconjugates, and FM1-43 (green) visualises membrane-associated structures. Images are shown without segmentation masks to preserve the original fluorescence and transmitted-light signals. Scale bars, 2.5 µm.

**Supplementary Information Table 8**: Strains used for unseen-organism analysis

| **Species** | **Code** | **Plate 1 strain** | **Relationship to training set** |
|---|---|---|---|
| *Haemophilus influenzae* | HFLU | L160570 | Unseen species |
| *Enterococcus faecalis* | EF | L127711 | Unseen species |
| *Enterococcus faecium* | EFM | L41428 | Unseen species |
| *Listeria monocytogenes* | LMON | L44679 | Unseen species |
| *Streptococcus agalactiae* | BSB | L135036 | Unseen species |

#### 7.1 Forced species classification and AST-routing assignments

Because the classifier was trained to distinguish only the 13 known SC-MAPS species, it did not contain an explicit unknown output. Consequently, every unseen cell received a probability distribution across the 13 learned species classes, regardless of whether its true organism was represented during training.

For each unseen organism, the predicted 13-class probability vectors were averaged across all available cells to obtain a mean forced species-probability distribution. These distributions were used to examine which learned species representations received the greatest probability when the classifier encountered unfamiliar organisms (Figure 4A). The assignments were interpreted as model preferences among learned image-derived phenotypes rather than valid species identifications.

The same species-probability distributions were additionally pooled into the eight predefined AST-routing groups by summing probabilities assigned to constituent species within each group. Mean routing-group probabilities were calculated separately for each unseen organism, allowing the distribution of predictions across the learned AST-routing categories to be examined (Figure 2E). These forced routing assignments were descriptive and were not interpreted as validated susceptibility-testing pathways or clinically appropriate routing recommendations for organisms absent from model development.

#### 7.2 Single-cell out-of-distribution detection

To determine whether unseen organisms could be explicitly distinguished from known species, four complementary OOD detection methods were evaluated: maximum softmax probability (MSP), normalised prediction entropy, k-nearest-neighbour (kNN) feature distance and class-conditional Mahalanobis distance.

MSP and entropy assessed uncertainty in the classifier's 13-species probability predictions, whereas kNN and Mahalanobis distance assessed deviation from the learned feature representation. For feature-based analyses, penultimate-layer representations were standardised and reduced to 50 dimensions using principal-component analysis (PCA). The kNN novelty score was calculated as the mean Euclidean distance to the 25 nearest cells in the known-species reference set, while Mahalanobis distance was calculated relative to the feature distributions of the known species, accounting for their covariance structure. Lower maximum SoftMax probability, higher entropy and larger feature-space distances indicated greater evidence that a cell differed from the known reference.

OOD detection thresholds were calibrated using known-species data only, without incorporating any of the five unseen organisms during threshold selection. The primary operating threshold was selected to retain 95% of known calibration cells. Discrimination between known and unseen organisms was evaluated using the area under the receiver operating characteristic curve (AUROC), known-species retention and unseen-organism rejection rates. Rejection rates were additionally assessed separately for each unseen organism to examine variation in detection performance between species (Figure 4C–D).

#### 7.3 Population-level OOD detection

To investigate whether combining evidence from multiple cells improved recognition of unseen organisms, cell-level novelty scores were averaged across populations containing 1, 5, 10, 20 or 50 cells. For each population size, 200 repeated sub-samplings were evaluated, and the aggregated novelty score was compared with the calibrated OOD rejection threshold. Detection performance was assessed alongside retention of populations from known SC-MAPS species (Figure 4E).

This analysis examined whether combining cell-level evidence could improve the distinction between known and unseen populations without requiring the classifier to assign an unfamiliar organism to one of its existing species categories. Repeated sub-samplings were treated as evaluations of cell-selection variability rather than independent biological replicates.

Together, forced classification and explicit OOD scoring provided complementary assessments of model behaviour towards unseen organisms. Forced probability distributions characterised their relationships to learned species and AST-routing representations, whereas OOD detection evaluated whether those organisms could be rejected as outside the known reference. These analyses were exploratory and did not establish validated species identification or clinical AST-routing performance for unseen organisms.

Feature-space kNN distance provided the highest discrimination between known and unseen cells (AUROC = 0.944), with 77.8% unseen-cell rejection and 94.3% known-cell retention at the selected operating threshold. Rejection varied between unseen organisms, ranging from 71.4% for HFLU to 88.0% for LMON. Population-level detection increased from 79.7% with one cell to 99.8% with five cells and 100% with 10–20 cells, while known-cell retention remained approximately 94% (Figure 4C–E).

#### 8. Learned feature-space visualisation of known and unseen organisms

To examine the phenotypic structure learned by the four-channel DenseNet121 concatenation model, high-dimensional feature representations were extracted from the penultimate layer, immediately preceding the final species-classification layer. A class-balanced subset of 2,000 cells per species was randomly selected from the independent-strain holdout, comprising 26,000 cells across the 13 known SC-MAPS species. Feature vectors were reduced to 50 dimensions using principal-component analysis (PCA), followed by two-dimensional embedding using Uniform Manifold Approximation and Projection (UMAP). UMAP was implemented using 30 nearest neighbours, a minimum distance of 0.15, Euclidean distance and a fixed random seed of 42. The embedding was fitted using the 13 known species to establish the reference representation shown in Figure 3B.

##### 8.1 Projection of unseen organisms into the known-species reference

To visualise unseen organisms relative to the known-species reference, 2,000 cells per unseen organism were selected, comprising 10,000 additional cells. Their penultimate-layer feature vectors were transformed using the PCA model fitted to the known species and projected into the existing UMAP embedding without refitting. This preserved the known-species reference and allowed the unseen organisms to be visualised within the same feature-space representation (Figure 4B).

##### 8.2 Additional UMAP analyses

To further examine the organisation of learned features, two supplementary embeddings were generated independently of the fixed-reference projection. First, a joint UMAP was fitted using feature representations from both known and unseen organisms, with cells distinguished by Gram status and whether their species were represented during model development (Supplementary Figure S10, panel A). This provided an alternative view of the broader organisation of Gram-positive and Gram-negative phenotypes when both populations contributed to the embedding. Second, a separate UMAP was fitted exclusively using the five unseen organisms, without including known-species cells (Supplementary Figure S10, panel B). This allowed the organisation and overlap of unseen-organism feature representations to be examined independently of the known-species reference.

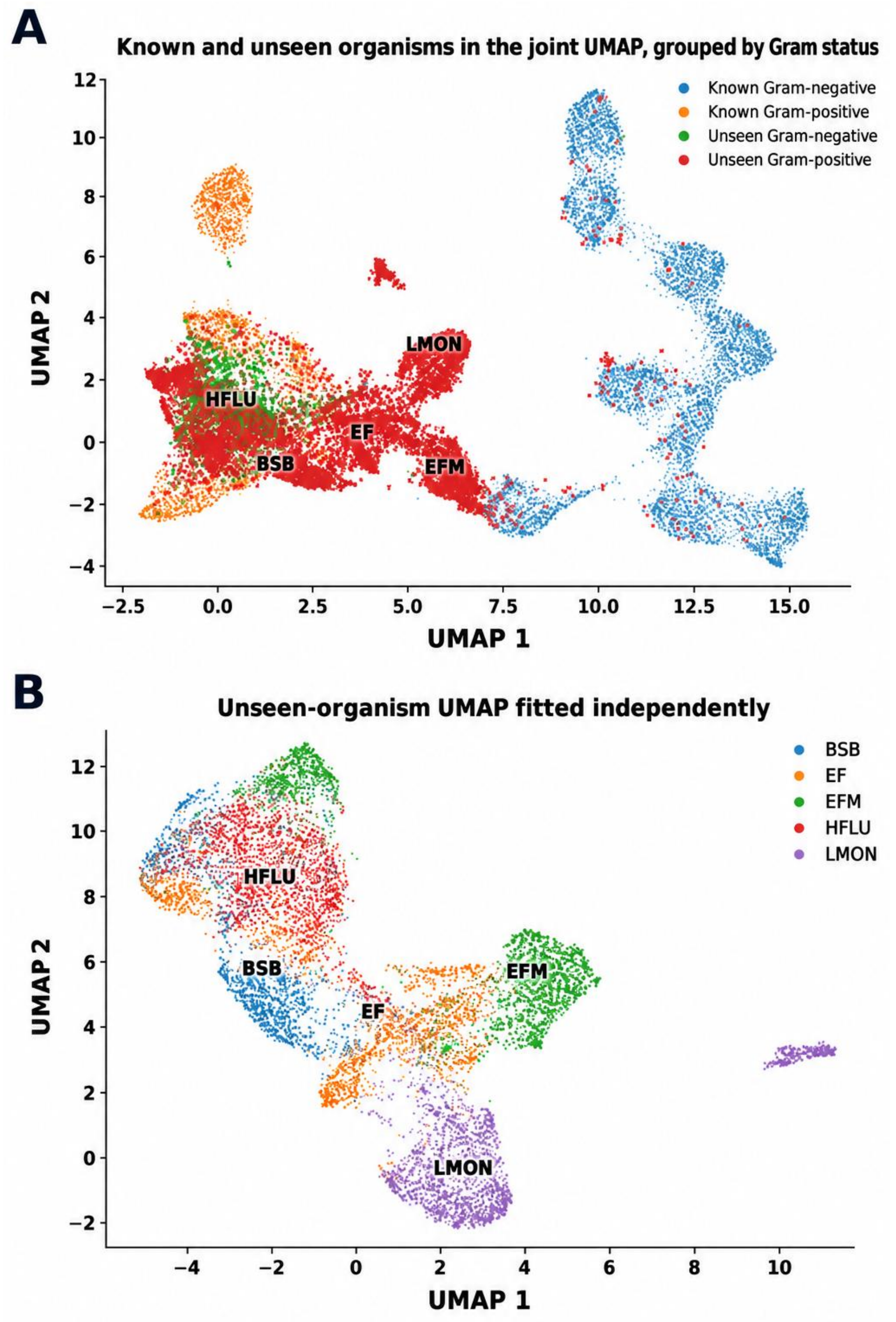

**Supplementary Figure S10: Alternative UMAP representations of known and unseen bacterial organisms.** Two-dimensional UMAP visualisations of penultimate-layer feature representations extracted from the four-channel DenseNet121 concatenation model. **(A)** Jointly fitted UMAP containing the 13 known SC-MAPS species and five unseen organisms, coloured according to Gram status and whether the species was represented during model development. **(B)** Independently fitted UMAP containing only the five unseen organisms: *Streptococcus agalactiae* (BSB), *Enterococcus faecalis* (EF), *Enterococcus faecium* (EFM), *Haemophilus influenzae* (HFLU) and *Listeria monocytogenes* (LMON), with each organism displayed in a distinct colour.

Both analyses used PCA-reduced feature representations and were performed independently of the fixed-reference UMAP shown in the main manuscript. The embeddings illustrate the organisation and overlap of learned phenotypic representations and were interpreted qualitatively. Coordinates and distances are not directly comparable between independently fitted UMAP embeddings and should not be interpreted as quantitative measures of biological or evolutionary relatedness.

### 9. Synthetic mixed species analysis

#### 9.1 Generation and evaluation of synthetic mixed-species populations

Synthetic two-species populations were generated using cell-level predictions from the independently held-out Plate 1 strains. All 78 pairwise combinations of the 13 SC-MAPS species were evaluated, using up to ten source-sample combinations per species pair. Populations contained 5, 10, 20, 50, 100 or 200 cells at target ratios of 50:50, 75:25, 90:10 and 95:5. Each species was evaluated as the dominant organism in asymmetric mixtures. Cells were sampled without replacement within each population, with 200 repeated sub-samplings per condition. Species-probability vectors were averaged across each population, and the two highest-probability species were selected as the aggregate predictions. Recovery was assessed for both constituent species and separately for dominant and minority organisms. Minority recovery was evaluated using 200-cell populations, while dominant recovery was assessed using 20- and 200-cell populations. The procedure is illustrated in Supplementary Figure S11.

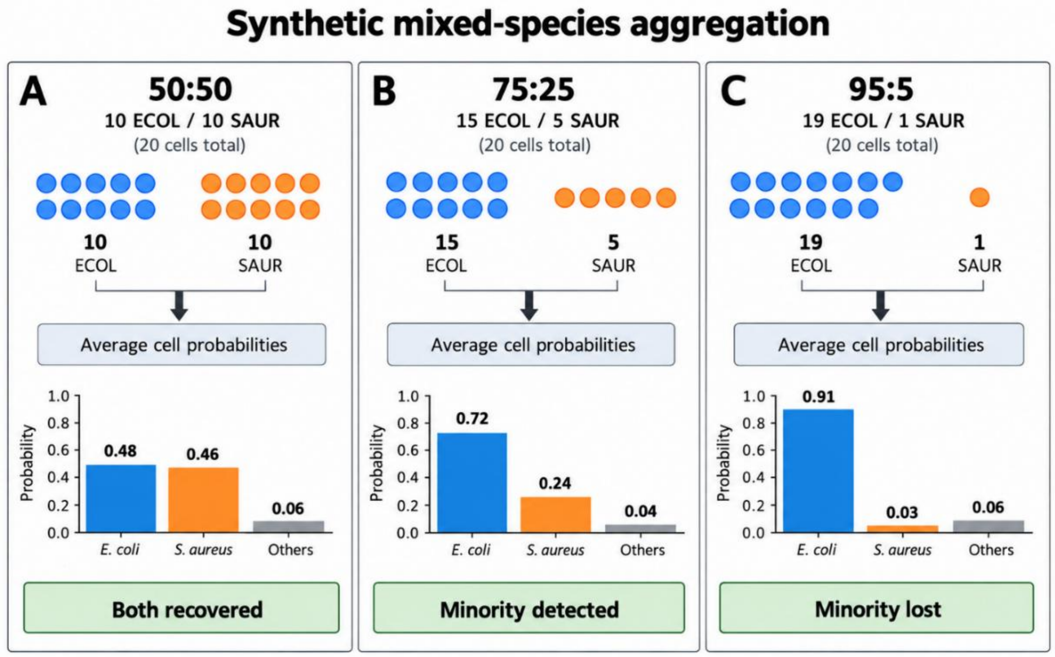

**Supplementary Figure S11: Illustrative examples of synthetic mixed-species aggregation.** Schematic examples showing how synthetic two-species populations were generated and analysed using Plate 1 holdout cells from *Escherichia coli* (ECOL) and *Staphylococcus aureus* (SAUR). For each example, cell-level 13-class species-probability vectors from the selected cells were averaged to generate a single population-level probability distribution, and the top-two aggregate species probabilities were used to assess recovery of the constituent organisms. **(A)** In a balanced 50:50 mixture (10 ECOL, 10 SAUR; 20 cells total), both species were recovered among the top-two aggregate predictions. **(B)** In a 75:25 mixture (15 ECOL, 5 SAUR), the minority species remained detectable in the top two predictions. **(C)** In a 95:5 mixture (19 ECOL, 1 SAUR), the minority species was no longer recovered among the top-two aggregate predictions. These illustrative examples show how population-level probability averaging preserves dominant-species identity while reducing detectability of low-abundance minority populations. The cell representations and plotted probability values are illustrative rather than measured observations from a particular mixture

##### 9.2 AST-routing recovery and randomisation analysis

For balanced 50:50 mixtures, aggregated species probabilities were pooled into the eight predefined AST-routing groups. Of the 78 species pairs, 72 belonged to different routing groups and six shared the same group. Recovery was assessed using the two highest-probability routing groups, or identification of the shared group where applicable. To assess whether improved routing recovery resulted simply from grouping species into fewer categories, 2,000 group-size-matched randomisations were performed, preserving the original routing-group sizes and keeping constituent species from different groups separate. For different-group mixtures, observed recovery of both routing groups was 95.4%, compared with a randomised mean of 78.9% (empirical $P = 0.0015$; Figure 2F). Same-group comparisons were evaluated descriptively

##### 9.3 Prediction heterogeneity using generalised Jensen–Shannon dispersion

Prediction heterogeneity was quantified using generalised Jensen–Shannon (JS) dispersion, calculated as the average Kullback–Leibler divergence between individual 13-class species-probability distributions and their population mean. Base-2 logarithms were used, expressing dispersion in bits. Lower values indicated greater agreement among cell predictions, whereas higher values indicated increasing heterogeneity.

Dispersion was evaluated in 20-cell pure and synthetic mixed populations across 200 repeated sub-samplings, with values averaged within each source-sample or mixture condition. Median JS dispersion was 0.00 bits for pure populations and 1.32 bits for mixed populations (Supplementary Figure S12). This demonstrated greater prediction heterogeneity in mixed populations, although JS dispersion was considered an exploratory measure rather than a validated method for detecting polymicrobial samples.

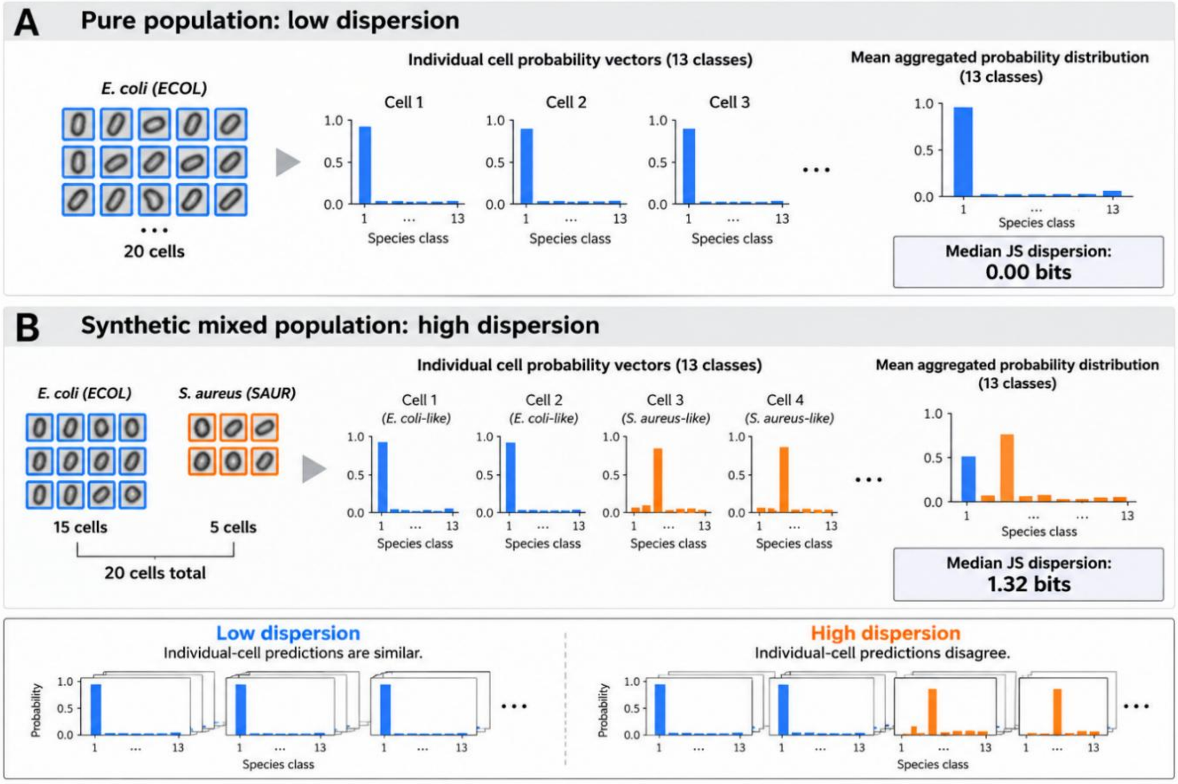

**Supplementary Figure S12: How prediction heterogeneity was quantified in pure and synthetic mixed populations. (A)** Schematic illustration of a pure 20-cell population derived from a single species, in which individual cells generate concordant 13-class species-probability distributions. Averaging these similar cell-level predictions produces a sharply peaked population-level probability distribution and low generalised Jensen–Shannon (JS) dispersion (median 0.00 bits). **(B)** Schematic illustration of a synthetic 20-cell mixed population containing cells from two species, in which individual cells generate heterogeneous probability distributions. Averaging these discordant cell-level predictions produces a broader population-level distribution and higher generalised JS dispersion (median 1.32 bits). This figure illustrates how dispersion captures disagreement among constituent cell predictions and therefore provides a measure of prediction heterogeneity within aggregated populations. Individual cell-level probability vectors are illustrative, whereas the reported median dispersion values are derived from the SC-MAPS analysis.

#### 10. Exploratory pixel-level attribution of multichannel species predictions

To investigate the image features contributing to bacterial species classification, we performed SmoothGrad–Integrated Gradients analysis on the DenseNet121 concatenation-fusion network. Unlike Grad-CAM, this approach provides pixel-level attribution at the original 96 × 96 input resolution, allowing the spatial contribution of individual imaging channels to be examined. We initially analysed 26 correctly classified cells from the independent holdout dataset, comprising two cells from each of the 13 species. Attribution was calculated relative to the highest-scoring alternative species and quantified separately for DAPI, phase contrast, WGA and FM1-43. Across the analysed cells, FM1-43 accounted for the largest proportion of total absolute attribution (40.8%), followed by DAPI (25.1%), phase contrast (23.2%) and WGA (10.8%). Spatial analysis further revealed that phase-contrast attribution was preferentially concentrated around bacterial boundaries, with 24 of 26 cells exhibiting greater attribution density within a two-pixel peripheral region than in the remaining cell interior. The mean boundary-to-interior attribution-density ratio was 1.44. These findings suggest that the network may exploit cell-boundary-associated contrast features alongside spatial patterns of fluorescence intensity when distinguishing bacterial species. However, given the limited number of cells examined, the selection of correctly classified examples and differences in the comparison species, these results remain exploratory. Attribution patterns indicate features associated with model predictions but do not independently establish their biological significance or causal contribution to classification.

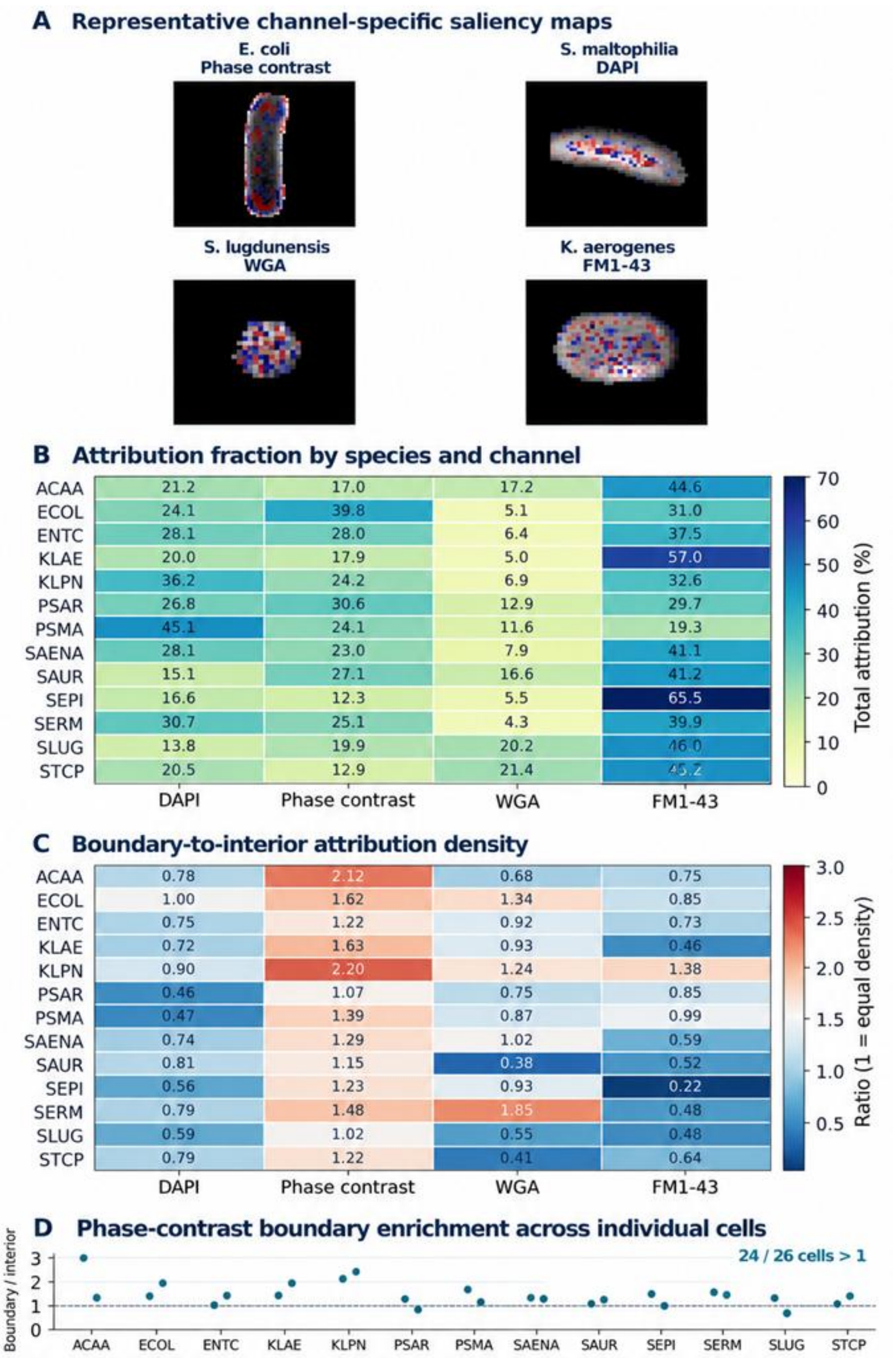

**Supplementary Figure S13 | Multichannel SmoothGrad–Integrated Gradients attribution analysis of bacterial classification.**: SmoothGrad–Integrated Gradients (SmoothGrad-IG) was applied to the four-channel DenseNet121 concatenation model using 26 correctly classified holdout cells (two per species) to investigate the image features contributing to species discrimination. **(A)** Representative channel-specific attribution maps for *E. coli* (phase contrast), *S. maltophilia* (DAPI), *S. lugdunensis* (WGA) and *K. aerogenes* (FM1-43). Red indicates positive and blue indicates negative contributions to the target-minus-rival species logit margin. **(B)** Relative attribution magnitude across the four imaging channels for each of the 13 bacterial species, expressed as a percentage of total attribution. **(C)** Boundary-to-interior attribution density ratios across species and channels. Values greater than 1 indicate higher attribution density at cell boundaries, defined as the outermost two pixels of the approximate foreground mask. **(D)** Phase-contrast boundary enrichment for individual cells, showing higher boundary than interior attribution density in 24 of 26 cells (92.3%). The dashed line indicates equal attribution density (ratio = 1).
